# Primary postpartum haemorrhage and longer-term physical, psychological, and psychosocial health outcomes for women and their partners: a mixed-methods systematic review

**DOI:** 10.1101/2022.08.24.22279096

**Authors:** Su Mon Latt, Fiona Alderdice, Madeline Elkington, Mahkawnghta Awng Shar, Jennifer J Kurinczuk, Rachel Rowe

**Author notes:** Corresponding author: (SL).

## Abstract

**Objectives:** Most research about outcomes following postpartum haemorrhage (PPH) has focused on immediate outcomes. There are fewer studies investigating longer-term maternal morbidity following PPH, resulting in a significant knowledge gap. This review aimed to synthesize the evidence about the longer-term physical and psychological consequences of primary PPH for women and their partners from high income settings.

**Methods:** The review was registered with PROSPERO and five electronic databases were searched. Studies were independently screened against the eligibility criteria by two reviewers and data were extracted from both quantitative and qualitative studies that reported non-immediate health outcomes of primary PPH.

**Results:** Data were included from 24 studies, of which 16 were quantitative, five were qualitative and three used mixed-methods. The included studies were of mixed methodological quality. Of the nine studies reporting outcomes beyond five years after birth, only two quantitative studies and one qualitative study had a follow-up period longer than ten years. Seven studies reported outcomes or experiences for partners. The evidence indicated that women with PPH were more likely to have persistent physical and psychological health problems after birth compared with women who did not have a PPH. These problems, including PTSD symptoms and cardiovascular disease, may be severe and extend for many years after birth and were more pronounced after a severe PPH, as indicated by a blood transfusion or hysterectomy. There was limited evidence about outcomes for partners after PPH, but conflicting evidence of association between PTSD and PPH among partners who witnessed PPH.

**Conclusion:** This review explored existing evidence about longer-term physical and psychological health outcomes among women who had a primary PPH in high income countries, and their partners. While the evidence about health outcomes beyond five years after PPH is limited, our findings indicate that women can experience long lasting negative impacts after primary PPH, including PTSD symptoms and cardiovascular disease, extending for many years after birth.

**PROSPERO registration number:** CRD42020161144

## Introduction

Postpartum haemorrhage (PPH) is defined by the World Health Organization (WHO) as blood loss of 500 ml or more from the genital tract within 24 hours after birth, (1) although the definition and the cut off point for the severity of PPH varies. (2) The incidence of PPH is increasing across the high-income countries. (3, 4) Most research about outcomes following PPH has focused on immediate outcomes including, for example, acute organ failure, (5) blood loss, hypovolemic shock and maternal death. (6, 7) There are fewer studies investigating longer-term maternal morbidity following PPH, resulting in a significant knowledge gap around the associations between PPH and subsequent health and wellbeing, including the impact on the psychosocial and emotional wellbeing of women and their partners. (5, 8–13).

A systematic review conducted in 2016 investigated the prevalence of women’s emotional and physical health problems following PPH and included six quantitative studies. (14) Several articles investigating potential physical or psychological consequences of PPH have been published since the last systematic search in April 2015. (15–17) A further systematic review found a potential association between PPH and post-traumatic stress disorder (PTSD), but this conclusion was limited by the small number of studies investigating this association. (18) Neither of these reviews included qualitative research exploring women’s experience of PPH, suggesting a comprehensive systematic review using an integrated mixed-methods approach would be valuable. (19) A mixed-methods systematic review was therefore conducted to synthesize the evidence about the longer-term physical and psychological consequences of PPH for women and their partners.

## Methods

The protocol for this review was registered with PROSPERO (CRD42020161144) and the review is reported according to the Preferred Reporting Items for Systematic Review and Meta-Analysis (PRISMA) 2020 checklist **(S1 Checklist**). (20)

### Eligibility criteria

We searched published literature for quantitative cohort, case-control, cross-sectional, or case series studies; qualitative studies including analysis of interviews, free text boxes/open-ended survey questions which explored women’s and partners’ experiences of care or seeking care and information following PPH, and/or their perceptions about the physical, psychological and psychosocial impact of PPH; and mixed-methods studies with an eligible qualitative or quantitative component. The inclusion/exclusion criteria in Table 1 were applied.

**Table 1.** Inclusion and exclusion criteria.

| Category | Inclusion criteria | Exclusion criteria |
| --- | --- | --- |
| **Population** | Women who gave birth after at least 24 weeks' gestation and had a PPH, and/or their partners. | Women who had a PPH and/or their partners who were a subset of study participants (e.g. women who had a PPH as a subset of women with a range of maternal morbidities) unless separate data from women who had a PPH could be retrieved. |
| **Exposure** | For quantitative studies, primary PPH, defined as:<br>Bleeding from the genital tract, within 24 hours of giving birth, with an estimated blood loss of >500 ml and/or blood transfusion within 24 hours of giving birth and/or having emergency peripartum hysterectomy (EPH) or arterial embolization following PPH, and/or peripartum fall in haemoglobin concentration of > 2g/dl.<br>For qualitative studies, PPH regardless of definition. | Quantitative studies where PPH was not the primary exposure of interest or in which PPH was not well-defined and measured by one of the criteria described. |
| **Comparison** | Studies with or without a control or comparison group, comprising participants who did not experience a PPH. | None |
| **Outcomes** | For quantitative studies, any non-immediate and non-fatal physical, psychological and psychosocial health outcomes for women and their partners after PPH.<br>For qualitative studies, women's and/or partners' experiences of care and of seeking care around the time of PPH and during the postpartum period, and their perceptions about the physical, psychological and psychosocial impact of PPH. | For quantitative studies: immediate outcomes, including maternal death following PPH, acute organ failure, sepsis, shock, and with intervention outcomes such as hysterectomy, intensive care admission, blood transfusion during initial birth admission; pregnancy-related outcomes, including secondary PPH, or recurrence of PPH in subsequent pregnancies. |
| **Time** | For quantitative studies, any follow-up duration after hospital discharge following PPH.<br>For qualitative studies, all studies regardless of follow-up duration. | None |
| **Setting** | High-income countries as defined by the World Bank (21) at the time of study. | Low/middle-income countries. |
| **Language** | English language | Papers which were not written in English |

### Search strategy

Searches (**S1 File)** were conducted in Medline (Ovid), Embase, Web of Science, CINAHL and PsycINFO on 16^th^ December 2019 and updated on 6^th^ January 2021 and on 31^st^ May 2022, from the inception of each database to the search date. Reference lists from all included full text articles were hand searched.

### Screening and study selection

Search results were imported into the reference management software Endnote and duplicates were removed. (22) Search results were then imported into Covidence software for screening and data extraction. (23) Titles and abstracts of all articles identified were independently screened by SL and MS against the inclusion criteria. Full text screening was conducted independently by SL and ME using the above eligibility criteria. Any disagreements were resolved by consensus between reviewers, involving RR when necessary.

### Data extraction

Data extraction was performed independently by SL, and either MS or ME. Study characteristics and outcome information were extracted for both quantitative and qualitative studies using bespoke data extraction forms developed and pilot-tested for this review **(S2 File**). For qualitative studies, all the text and themes reported under findings or results from included studies were imported into NVivo software and treated as a primary textual data source for analysis. Data from mixed method studies were entered in both forms as appropriate.

### Risk of bias and quality assessment

Risk of bias and quality assessment for the included studies was conducted by SM, MA and ME, with two independent reviewers for each study, with any disagreements resolved by consensus as described above. Included quantitative studies (and the quantitative component of mixed-methods studies) were assessed using the risk of bias assessment for non-randomized studies (ROBANS) tool **(S1 Table).** (24)

Quality assessment for qualitative studies was conducted using the Critical Appraisal Skills Programme (CASP) tool. (25) The response options for Questions 1-9 were modified from yes/no/unclear to include the option to respond ‘partly’ where a paper addressed some items in the checklist associated with the question, but not all. Quantitative and qualitative components of mixed-methods studies were assessed and reported separately.

### Data synthesis

Since the quantitative studies were not homogeneous in terms of study design and outcome measurements, we used a narrative synthesis approach, presenting quantitative study results in structured tables. (26) Qualitative studies were synthesised using the approach described by Thomas and Harden. (27–29) This involved: 1) free line-by-line coding of the findings of primary studies; 2) the organization of codes in related areas to construct ‘descriptive’ themes; and 3) the development of ‘analytical’ themes to answer the research questions. (27–29)

We used the Joanna Briggs Institute (JBI) convergent segregated approach for mixed-methods systematic reviews, whereby following separate analysis of the quantitative and qualitative evidence, the similarities and differences in findings were compared to produce a narrative summary. (30)

## Results

The search yielded 15,742 references, of which 7,625 were duplicates and were removed (***Fig 1*)**. After removing duplicates, a total of 8,117 references remained. Following title and abstract screening, 127 references were included for full text review. An additional six references were identified through other sources. After removal of ineligible studies, 33 papers reporting 24 studies were included in the review, of which 16 were quantitative, five qualitative, and three mixed-methods studies.

**Fig 1.**
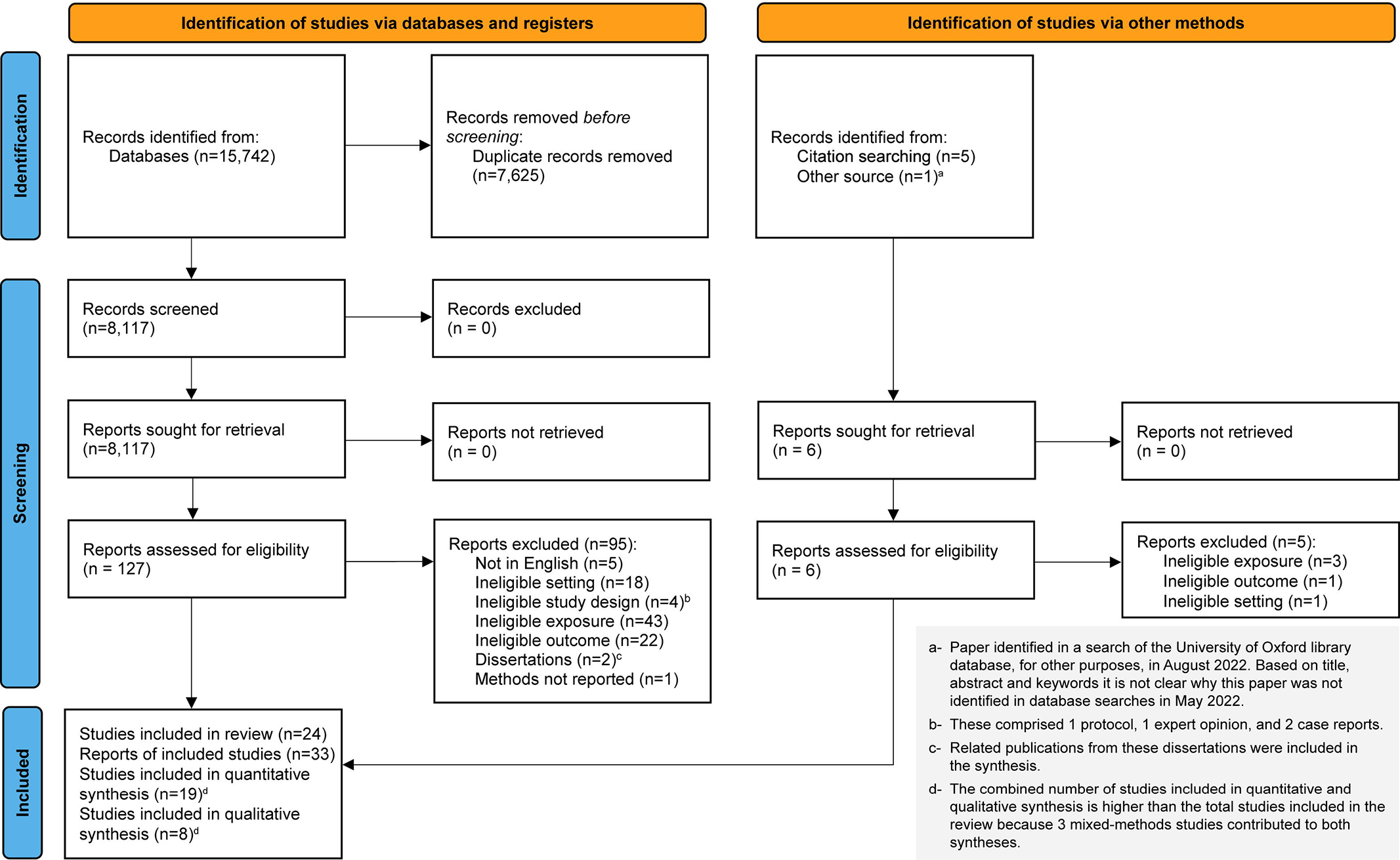
PRISMA flow diagram for study selection (20)

### Characteristics of included studies

Included studies were conducted in Australia, New Zealand, France, the Netherlands, Sweden, Switzerland, South Korea, Canada, the UK and the United States, between 2005 and 2021. The follow-up time was from hospital discharge following PPH to up to six weeks postpartum in seven studies, (31–38) up to a year in four studies, (12, 16, 39–42) up to five years in three studies, (43–46) and longer than five years in nine studies. (47–58) One qualitative study did not report on follow-up duration. (59) The definitions for PPH and severe PPH used in these studies varied widely. In studies defining PPH according to blood loss volume, this varied from 500ml to 1000ml for vaginal birth and from 750 ml to 1500 ml for Caesarean birth. In four studies, PPH was identified by using ICD 10 codes recorded in hospital or insurance databases. (35, 42, 51, 52) Definitions for severe PPH also varied widely, from >1000ml to >2000ml. In seven studies where the exposure was severe PPH, severity was also indicated by combining blood loss volume with interventions such as emergency peripartum hysterectomy (EPH) and/or arterial embolization following PPH. (39, 44, 47–49, 54, 56) PPH was not well defined in one qualitative study, (38) however, all qualitative studies were included regardless of their definitions as these were not part of the eligibility criteria for qualitative studies.

Summary characteristics of all included studies are presented in Table 2.

**Table 2.**
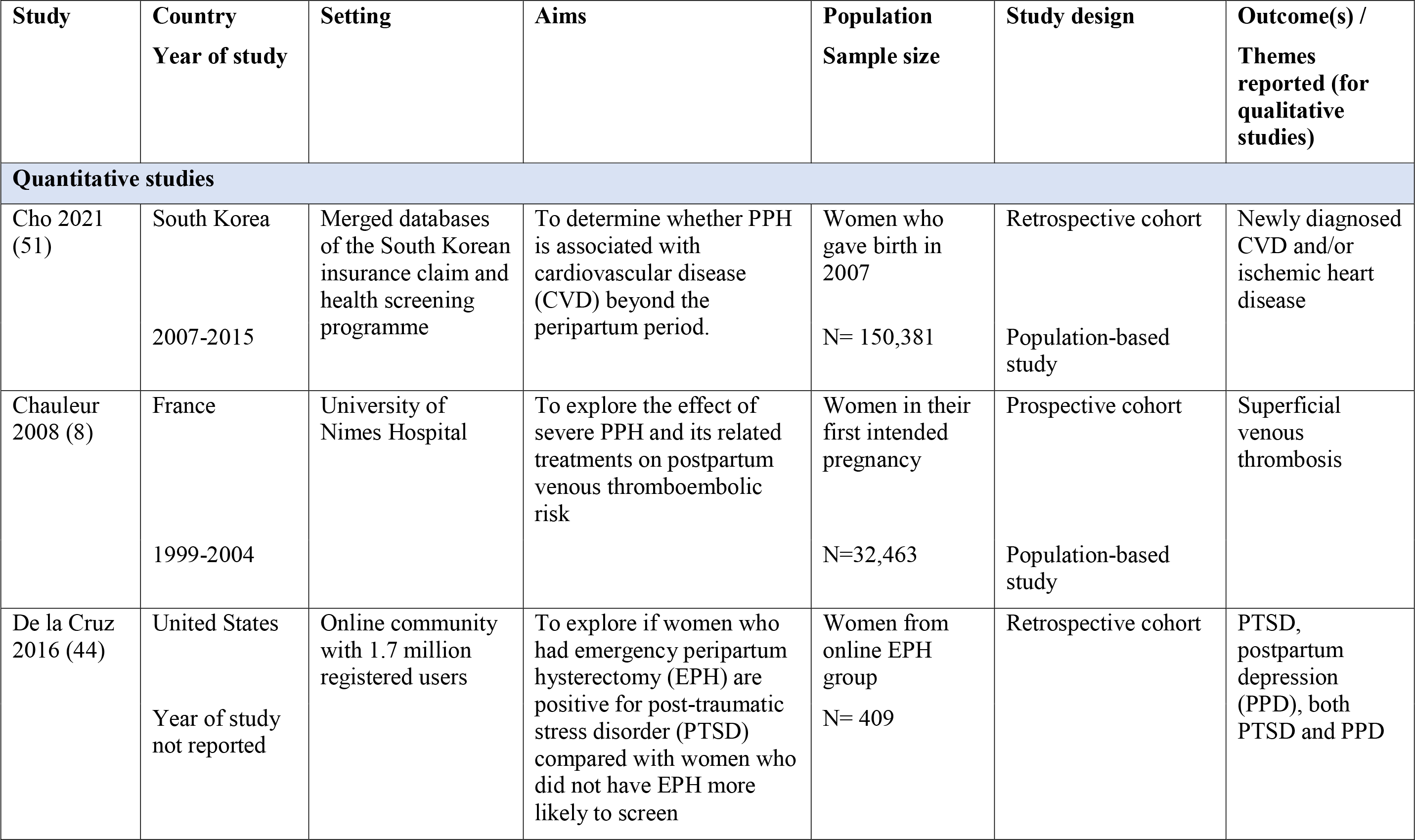
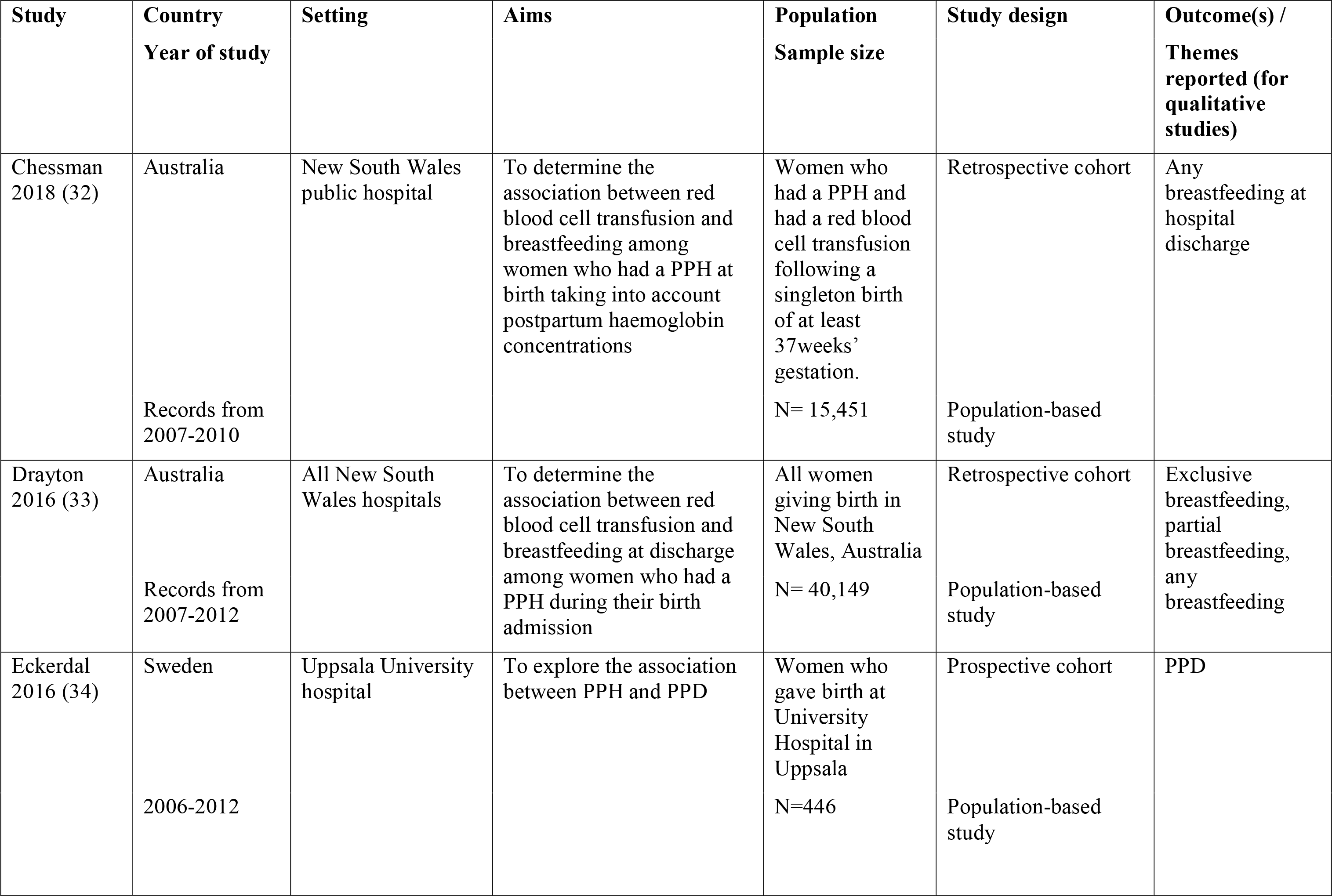
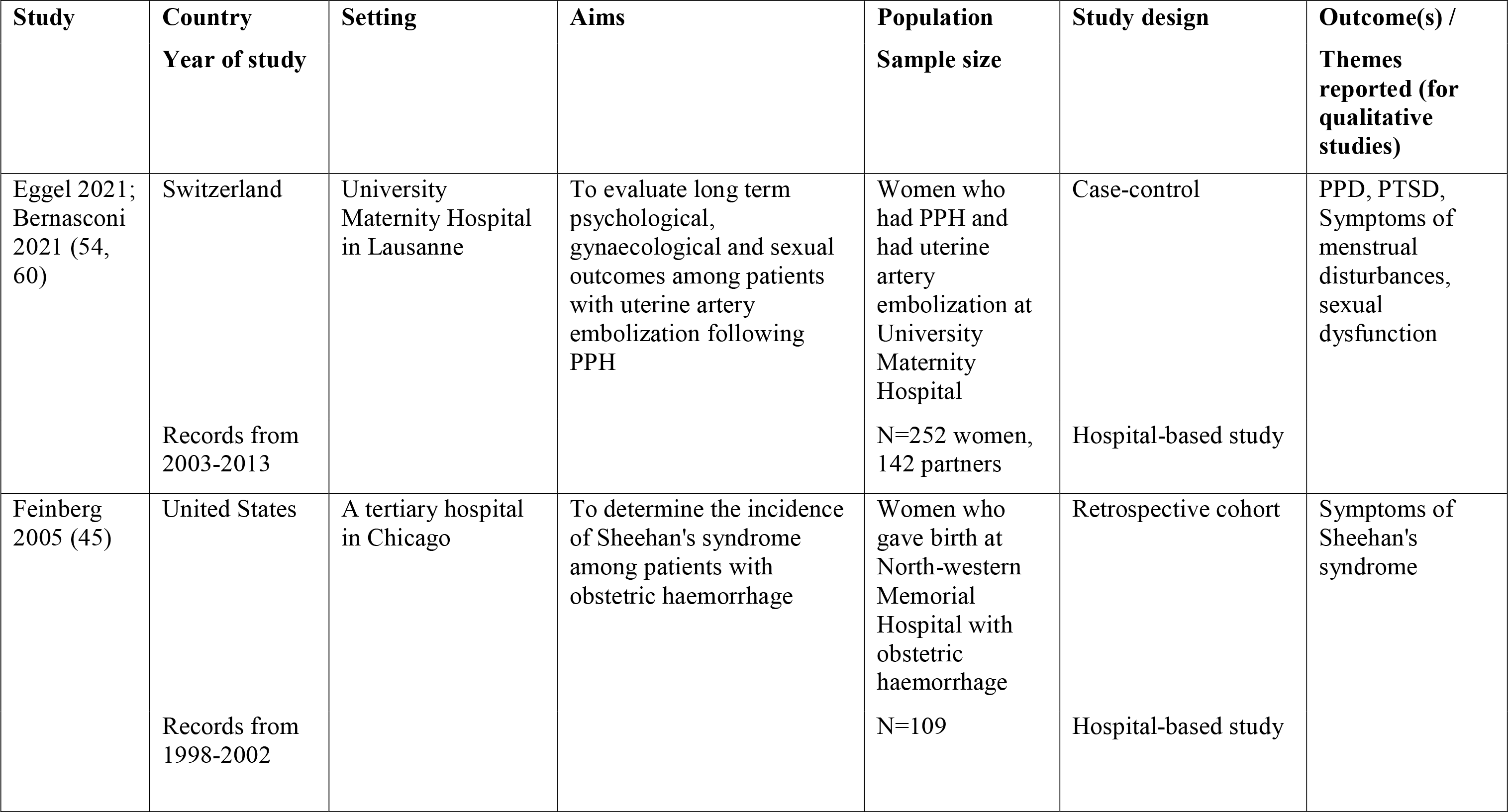
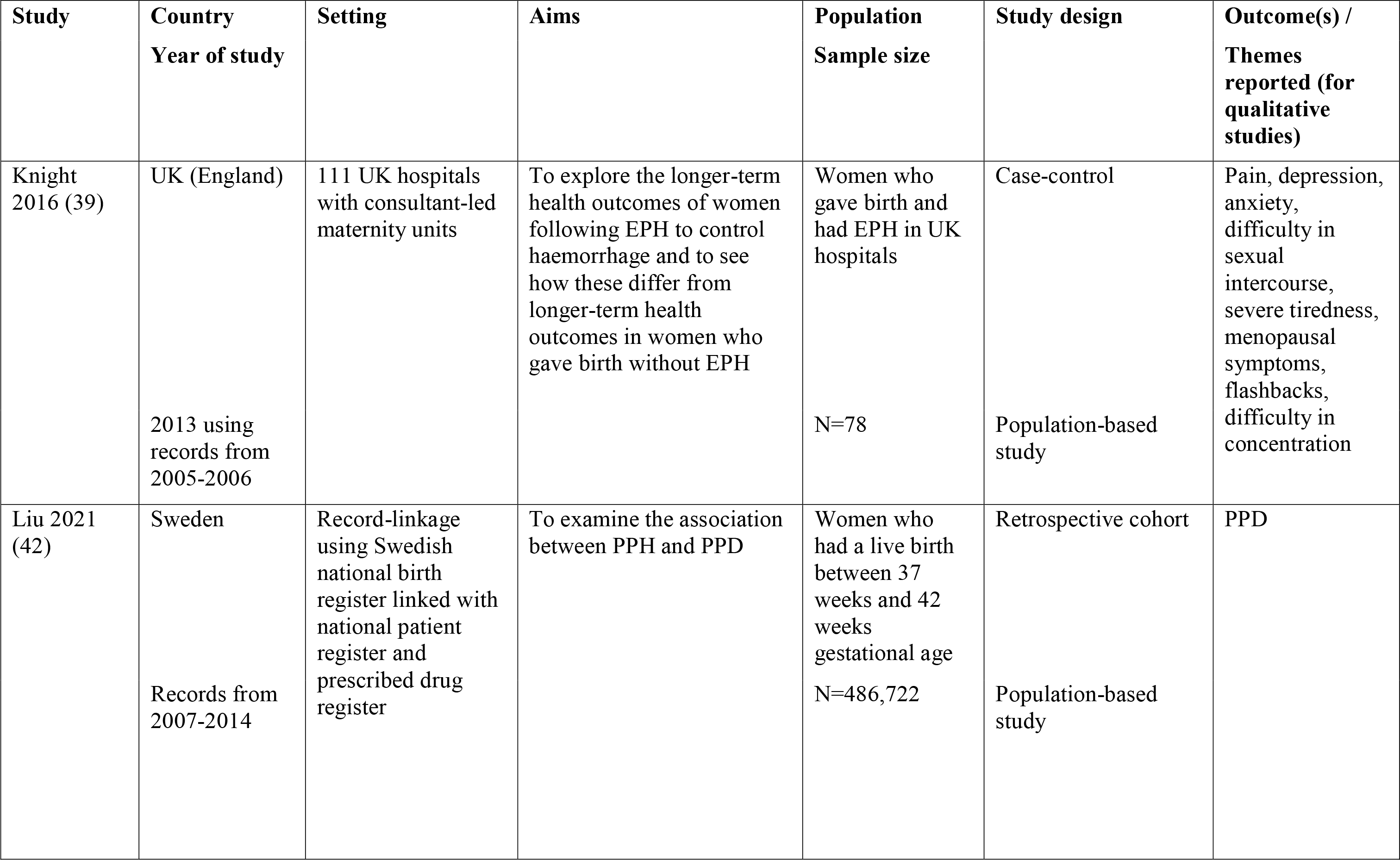
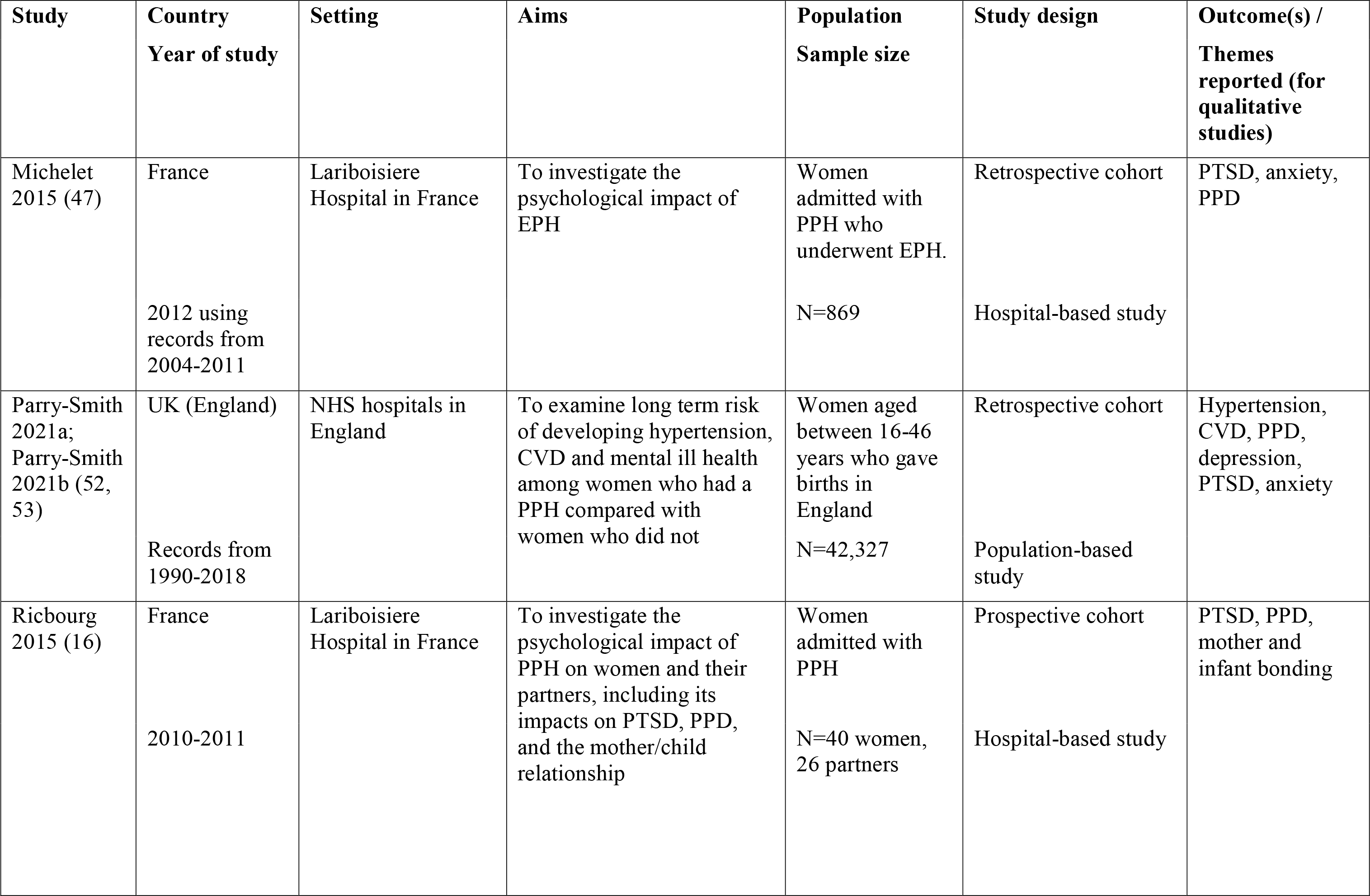
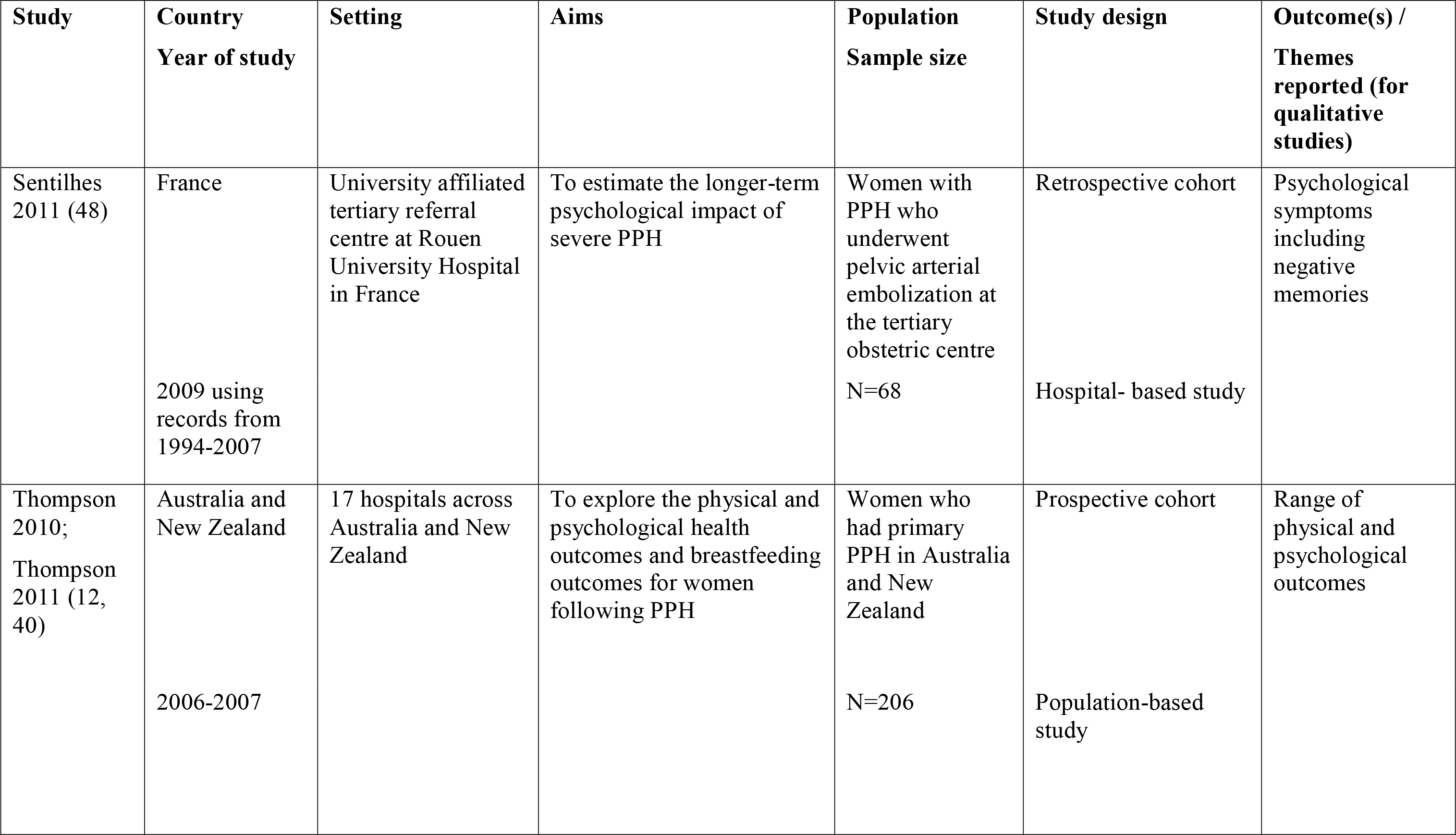
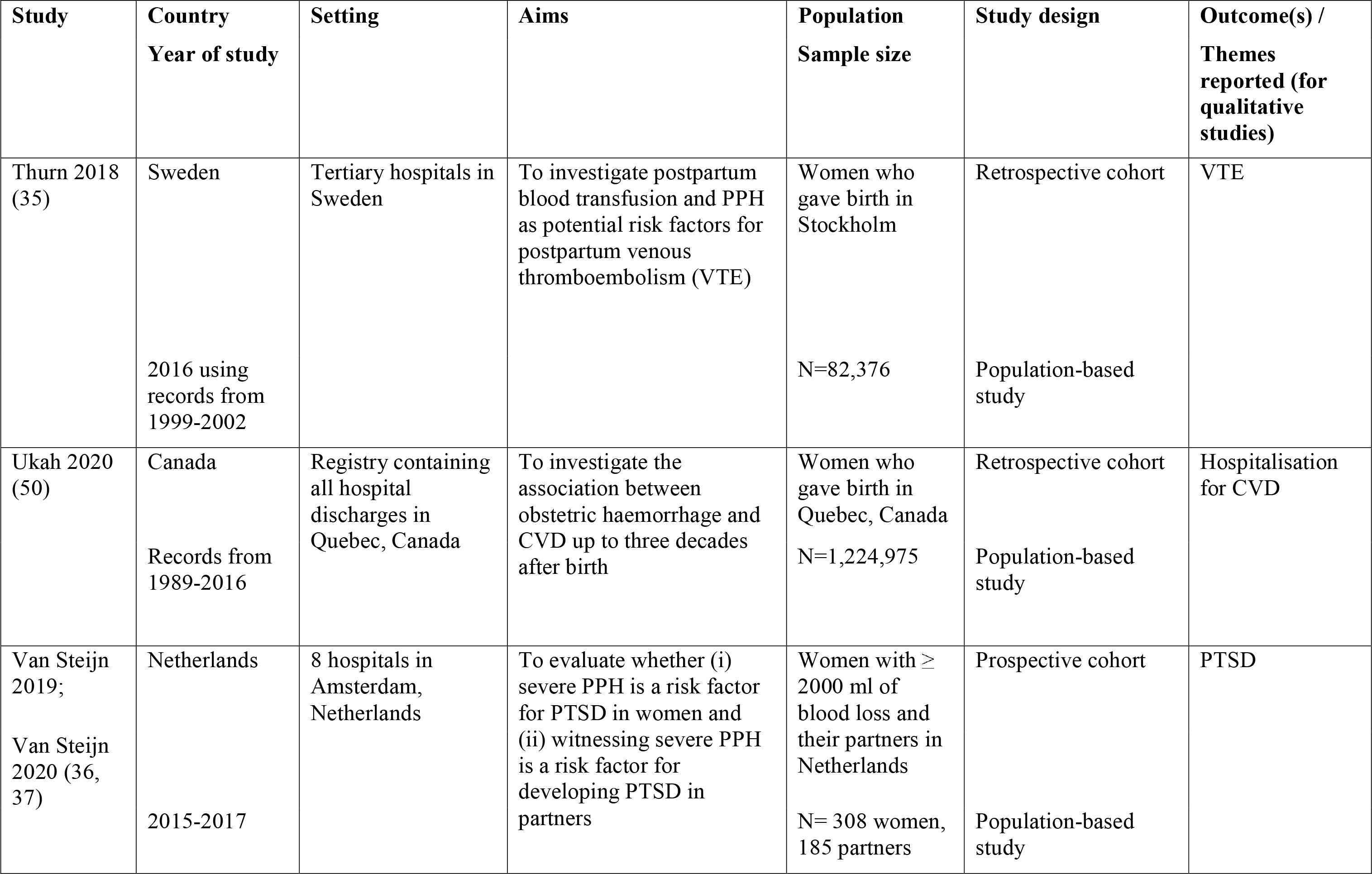
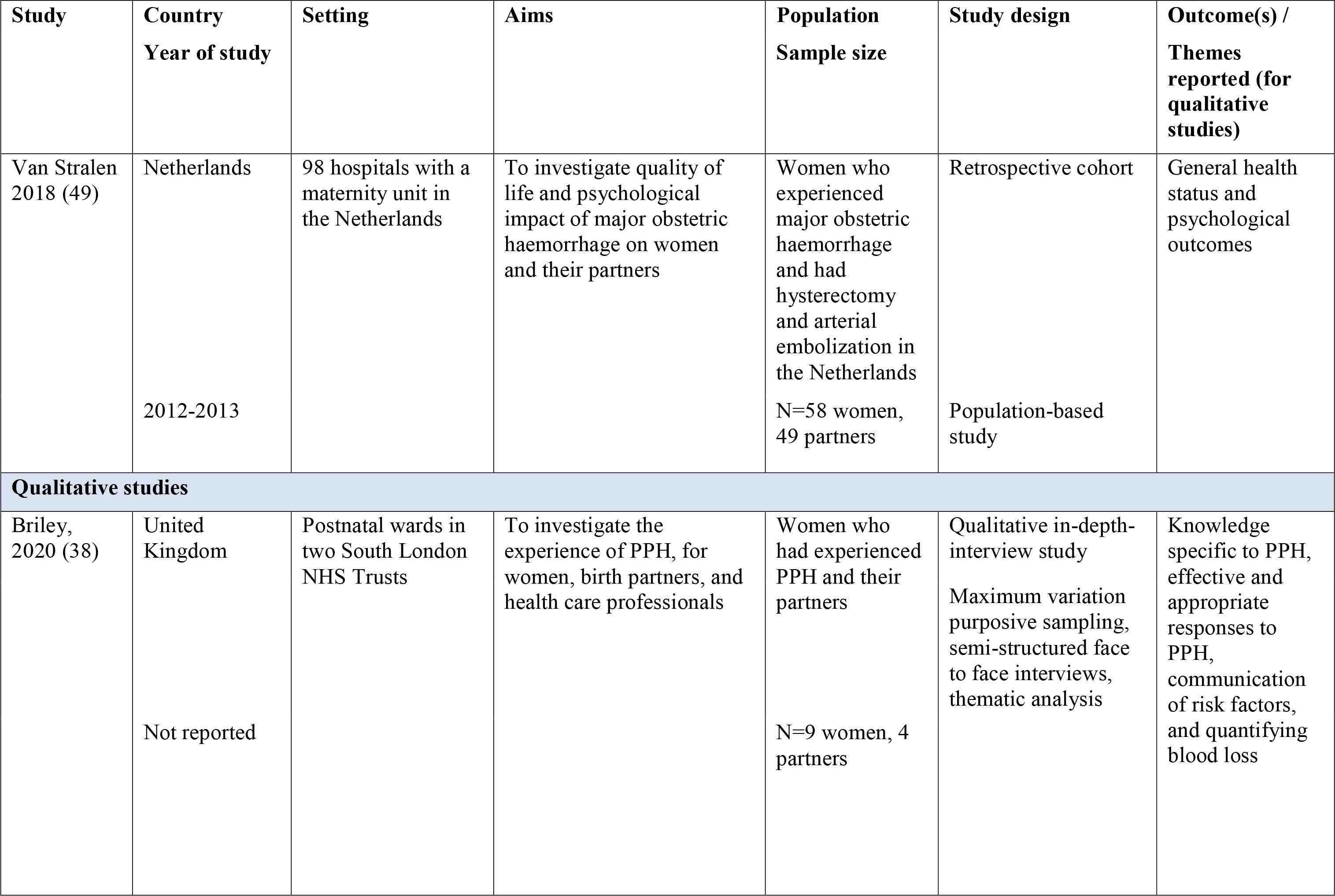
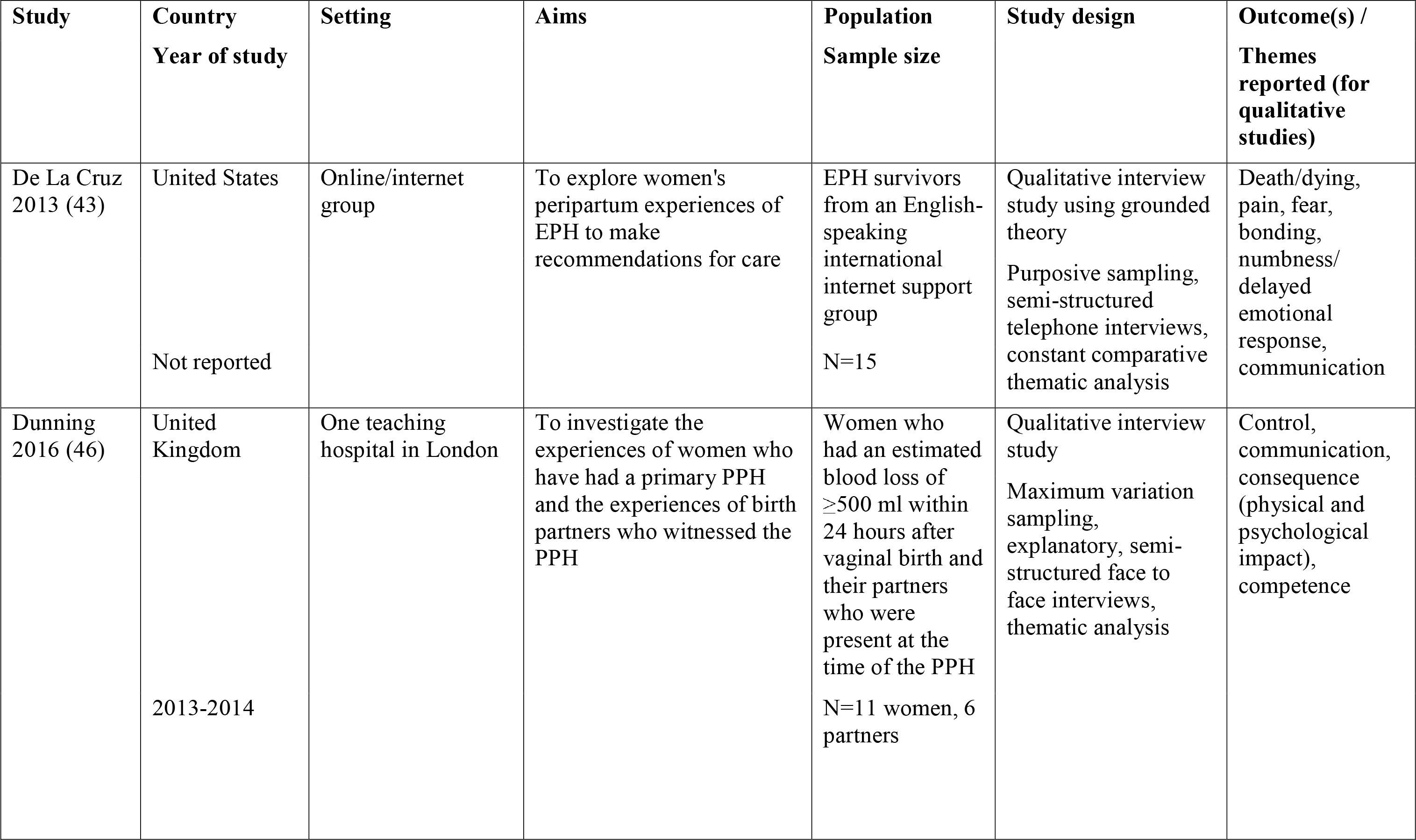
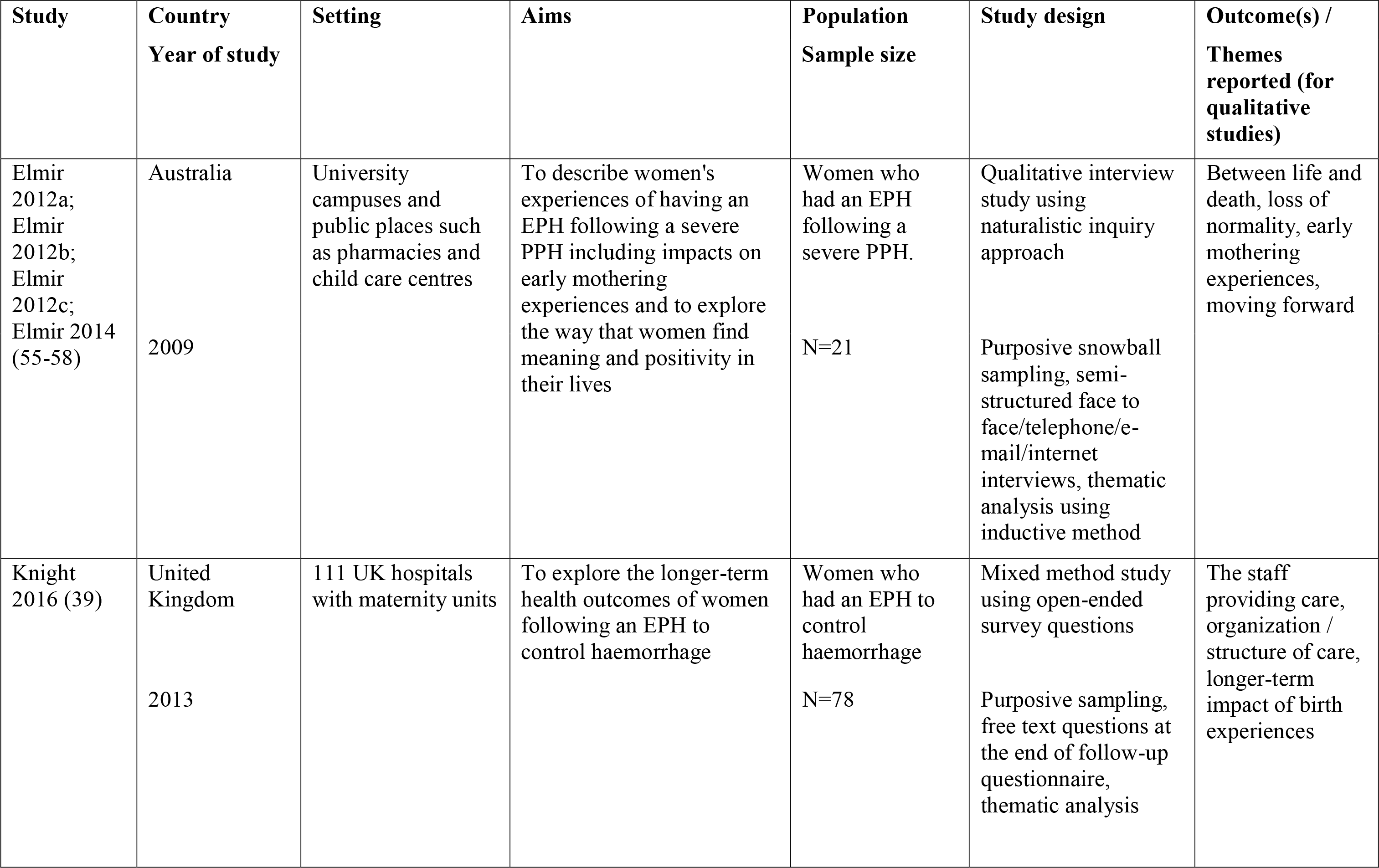
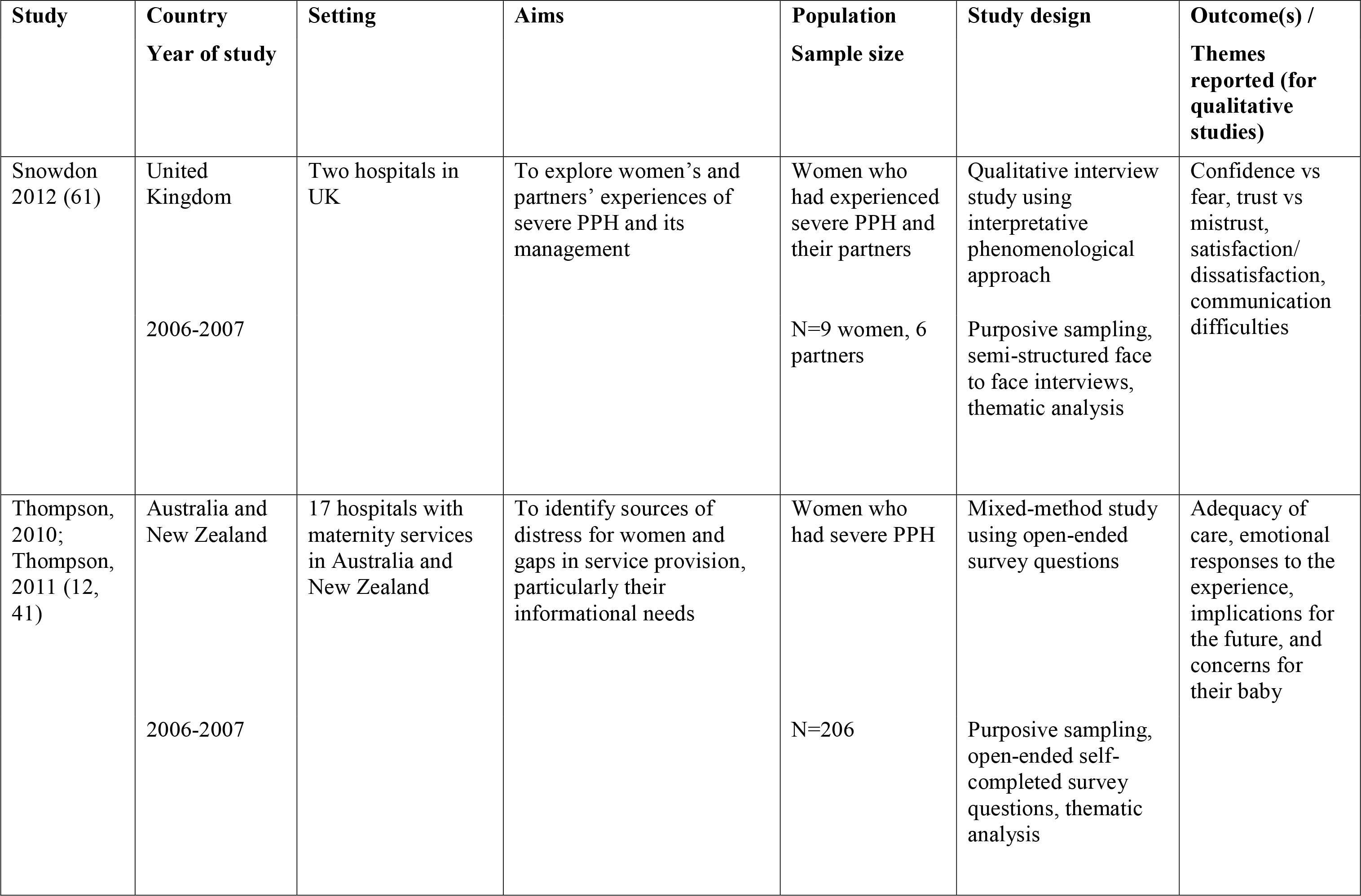
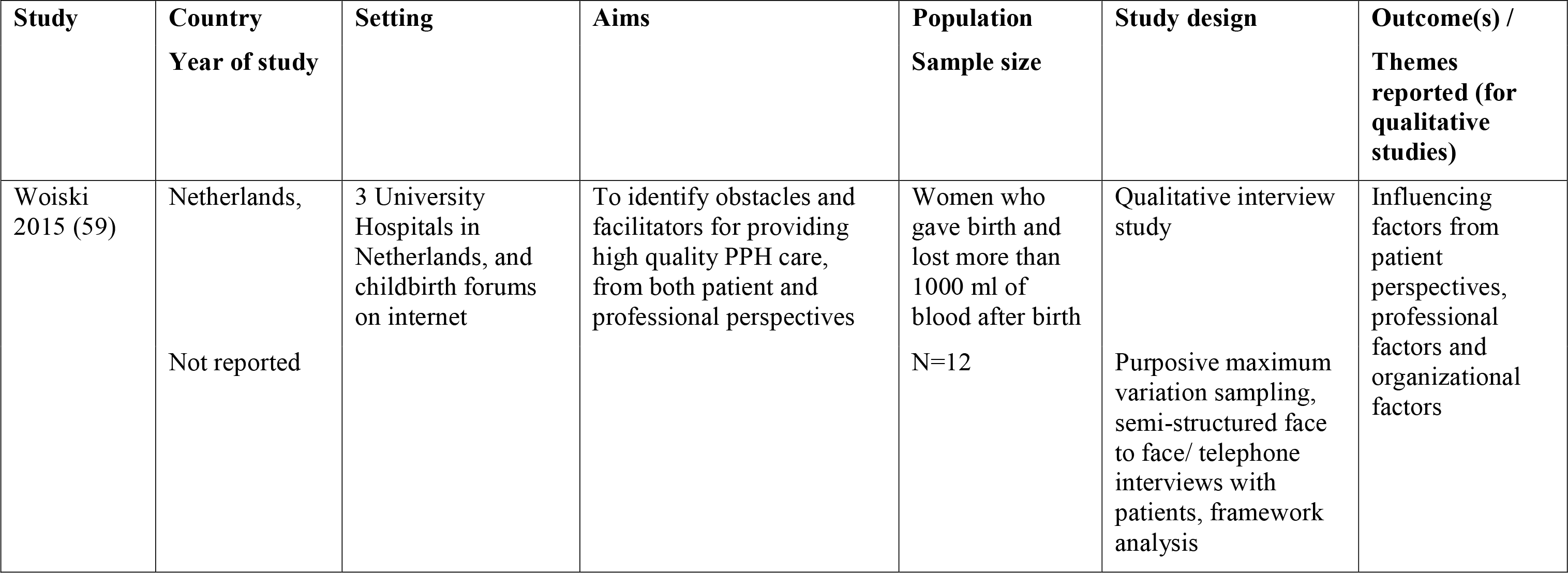
Characteristics of included studies.

| **Study** | **Country**<br>**Year of study** | **Setting** | **Aims** | **Population**<br>**Sample size** | **Study design** | **Outcome(s) /**<br>**Themes**<br>**reported (for**<br>**qualitative**<br>**studies)** |
| --- | --- | --- | --- | --- | --- | --- |
| **Quantitative studies** |  |  |  |  |  |  |
| Cho 2021<br>(51) | South Korea<br>2007-2015 | Merged databases<br>of the South Korean<br>insurance claim and<br>health screening<br>programme | To determine whether PPH<br>is associated with<br>cardiovascular disease<br>(CVD) beyond the<br>peripartum period. | Women who<br>gave birth in<br>2007<br>N= 150,381 | Retrospective cohort<br>Population-based<br>study | Newly diagnosed<br>CVD and/or<br>ischemic heart<br>disease |
| Chauleur<br>2008 (8) | France<br>1999-2004 | University of<br>Nimes Hospital | To explore the effect of<br>severe PPH and its related<br>treatments on postpartum<br>venous thromboembolic<br>risk | Women in their<br>first intended<br>pregnancy<br>N=32,463 | Prospective cohort<br>Population-based<br>study | Superficial<br>venous<br>thrombosis |
| De la Cruz<br>2016 (44) | United States<br>Year of study<br>not reported | Online community<br>with 1.7 million<br>registered users | To explore if women who<br>had emergency peripartum<br>hysterectomy (EPH) are<br>positive for post-traumatic<br>stress disorder (PTSD)<br>compared with women who<br>did not have EPH more<br>likely to screen | Women from<br>online EPH<br>group<br>N= 409 | Retrospective cohort | PTSD,<br>postpartum<br>depression<br>(PPD), both<br>PTSD and PPD |
| Chessman<br>2018 (32) | Australia<br>Records from<br>2007-2010 | New South Wales<br>public hospital | To determine the<br>association between red<br>blood cell transfusion and<br>breastfeeding among<br>women who had a PPH at<br>birth taking into account<br>postpartum haemoglobin<br>concentrations | Women who<br>had a PPH and<br>had a red blood<br>cell transfusion<br>following a<br>singleton birth<br>of at least<br>37weeks' gestation.<br>N= 15,451 | Retrospective cohort<br>Population-based<br>study | Any<br>breastfeeding at<br>hospital<br>discharge |
| Drayton<br>2016 (33) | Australia<br>Records from<br>2007-2012 | All New South<br>Wales hospitals | To determine the<br>association between red<br>blood cell transfusion and<br>breastfeeding at discharge<br>among women who had a<br>PPH during their birth<br>admission | All women<br>giving birth in<br>New South<br>Wales, Australia<br>N= 40,149 | Retrospective cohort<br>Population-based<br>study | Exclusive<br>breastfeeding,<br>partial<br>breastfeeding,<br>any<br>breastfeeding |
| Eckerdal<br>2016 (34) | Sweden<br>2006-2012 | Uppsala University<br>hospital | To explore the association<br>between PPH and PPD | Women who<br>gave birth at<br>University<br>Hospital in<br>Uppsala<br>N=446 | Prospective cohort<br>Population-based<br>study | PPD |
| Eggel 2021;<br>Bernasconi<br>2021 (54,<br>60) | Switzerland<br>Records from<br>2003-2013 | University<br>Maternity Hospital<br>in Lausanne | To evaluate long term<br>psychological,<br>gynaecological and sexual<br>outcomes among patients<br>with uterine artery<br>embolization following<br>PPH | Women who<br>had PPH and<br>had uterine<br>artery<br>embolization at<br>University<br>Maternity<br>Hospital<br>N=252 women,<br>142 partners | Case-control<br>Hospital-based study | PPD, PTSD,<br>Symptoms of<br>menstrual<br>disturbances,<br>sexual<br>dysfunction |
| Feinberg<br>2005 (45) | United States<br>Records from<br>1998-2002 | A tertiary hospital<br>in Chicago | To determine the incidence<br>of Sheehan's syndrome<br>among patients with<br>obstetric haemorrhage | Women who<br>gave birth at<br>North-western<br>Memorial<br>Hospital with<br>obstetric<br>haemorrhage<br>N=109 | Retrospective cohort<br>Hospital-based study | Symptoms of<br>Sheehan's<br>syndrome |
| Knight<br>2016 (39) | UK (England)<br>2013 using<br>records from<br>2005-2006 | 111 UK hospitals<br>with consultant-led<br>maternity units | To explore the longer-term<br>health outcomes of women<br>following EPH to control<br>haemorrhage and to see<br>how these differ from<br>longer-term health<br>outcomes in women who<br>gave birth without EPH | Women who<br>gave birth and<br>had EPH in UK<br>hospitals<br>N=78 | Case-control<br>Population-based<br>study | Pain, depression,<br>anxiety,<br>difficulty in<br>sexual<br>intercourse,<br>severe tiredness,<br>menopausal<br>symptoms,<br>flashbacks,<br>difficulty in<br>concentration |
| Liu 2021<br>(42) | Sweden<br>Records from<br>2007-2014 | Record-linkage<br>using Swedish<br>national birth<br>register linked with<br>national patient<br>register and<br>prescribed drug<br>register | To examine the association<br>between PPH and PPD | Women who<br>had a live birth<br>between 37<br>weeks and 42<br>weeks<br>gestational age<br>N=486,722 | Retrospective cohort<br>Population-based<br>study | PPD |
| Michelet<br>2015 (47) | France<br>2012 using<br>records from<br>2004-2011 | Lariboisiere<br>Hospital in France | To investigate the<br>psychological impact of<br>EPH | Women<br>admitted with<br>PPH who<br>underwent EPH.<br>N=869 | Retrospective cohort<br>Hospital-based study | PTSD, anxiety,<br>PPD |
| Parry-Smith<br>2021a;<br>Parry-Smith<br>2021b (52,<br>53) | UK (England)<br>Records from<br>1990-2018 | NHS hospitals in<br>England | To examine long term risk<br>of developing hypertension,<br>CVD and mental ill health<br>among women who had a<br>PPH compared with<br>women who did not | Women aged<br>between 16-46<br>years who gave<br>births in<br>England<br>N=42,327 | Retrospective cohort<br>Population-based<br>study | Hypertension,<br>CVD, PPD,<br>depression,<br>PTSD, anxiety |
| Ricbourg<br>2015 (16) | France<br>2010-2011 | Lariboisiere<br>Hospital in France | To investigate the<br>psychological impact of<br>PPH on women and their<br>partners, including its<br>impacts on PTSD, PPD,<br>and the mother/child<br>relationship | Women<br>admitted with<br>PPH<br>N=40 women,<br>26 partners | Prospective cohort<br>Hospital-based study | PTSD, PPD,<br>mother and<br>infant bonding |
| Sentilhes<br>2011 (48) | France<br>2009 using<br>records from<br>1994-2007 | University affiliated<br>tertiary referral<br>centre at Rouen<br>University Hospital<br>in France | To estimate the longer-term<br>psychological impact of<br>severe PPH | Women with<br>PPH who<br>underwent<br>pelvic arterial<br>embolization at<br>the tertiary<br>obstetric centre<br>N=68 | Retrospective cohort<br>Hospital- based study | Psychological<br>symptoms<br>including<br>negative<br>memories |
| Thompson<br>2010;<br>Thompson<br>2011 (12,<br>40) | Australia and<br>New Zealand<br>2006-2007 | 17 hospitals across<br>Australia and New<br>Zealand | To explore the physical and<br>psychological health<br>outcomes and breastfeeding<br>outcomes for women<br>following PPH | Women who<br>had primary<br>PPH in Australia<br>and New<br>Zealand<br>N=206 | Prospective cohort<br>Population-based<br>study | Range of<br>physical and<br>psychological<br>outcomes |
| Thurn 2018<br>(35) | Sweden<br>2016 using<br>records from<br>1999-2002 | Tertiary hospitals in<br>Sweden | To investigate postpartum<br>blood transfusion and PPH<br>as potential risk factors for<br>postpartum venous<br>thromboembolism (VTE) | Women who<br>gave birth in<br>Stockholm<br>N=82,376 | Retrospective cohort<br>Population-based<br>study | VTE |
| Ukah 2020<br>(50) | Canada<br>Records from<br>1989-2016 | Registry containing<br>all hospital<br>discharges in<br>Quebec, Canada | To investigate the<br>association between<br>obstetric haemorrhage and<br>CVD up to three decades<br>after birth | Women who<br>gave birth in<br>Quebec, Canada<br>N=1,224,975 | Retrospective cohort<br>Population-based<br>study | Hospitalisation<br>for CVD |
| Van Steijn<br>2019;<br>Van Steijn<br>2020 (36,<br>37) | Netherlands<br>2015-2017 | 8 hospitals in<br>Amsterdam,<br>Netherlands | To evaluate whether (i)<br>severe PPH is a risk factor<br>for PTSD in women and<br>(ii) witnessing severe PPH<br>is a risk factor for<br>developing PTSD in<br>partners | Women with ≥<br>2000 ml of<br>blood loss and<br>their partners in<br>Netherlands<br>N= 308 women,<br>185 partners | Prospective cohort<br>Population-based<br>study | PTSD |

| Study | Country<br>Year of study | Setting | Aims | Population<br>Sample size | Study design | Outcome(s) /<br>Themes<br>reported (for<br>qualitative<br>studies) |
| --- | --- | --- | --- | --- | --- | --- |
| Van Stralen<br>2018 (49) | Netherlands<br>2012-2013 | 98 hospitals with a<br>maternity unit in<br>the Netherlands | To investigate quality of<br>life and psychological<br>impact of major obstetric<br>haemorrhage on women<br>and their partners | Women who<br>experienced<br>major obstetric<br>haemorrhage<br>and had<br>hysterectomy<br>and arterial<br>embolization in<br>the Netherlands<br>N=58 women,<br>49 partners | Retrospective cohort<br>Population-based<br>study | General health<br>status and<br>psychological<br>outcomes |
| **Qualitative studies** |  |  |  |  |  |  |
| Briley,<br>2020 (38) | United<br>Kingdom<br>Not reported | Postnatal wards in<br>two South London<br>NHS Trusts | To investigate the<br>experience of PPH, for<br>women, birth partners, and<br>health care professionals | Women who<br>had experienced<br>PPH and their<br>partners<br>N=9 women, 4<br>partners | Qualitative in-depth-<br>interview study<br>Maximum variation<br>purposive sampling,<br>semi-structured face<br>to face interviews,<br>thematic analysis | Knowledge<br>specific to PPH,<br>effective and<br>appropriate<br>responses to<br>PPH,<br>communication<br>of risk factors,<br>and quantifying<br>blood loss |
| De La Cruz<br>2013 (43) | United States<br>Not reported | Online/internet<br>group | To explore women's<br>peripartum experiences of<br>EPH to make<br>recommendations for care | EPH survivors<br>from an English-<br>speaking<br>international<br>internet support<br>group<br>N=15 | Qualitative interview<br>study using grounded<br>theory<br>Purposive sampling,<br>semi-structured<br>telephone interviews,<br>constant comparative<br>thematic analysis | Death/dying,<br>pain, fear,<br>bonding,<br>numbness/<br>delayed<br>emotional<br>response,<br>communication |
| Dunning<br>2016 (46) | United<br>Kingdom<br>2013-2014 | One teaching<br>hospital in London | To investigate the<br>experiences of women who<br>have had a primary PPH<br>and the experiences of birth<br>partners who witnessed the<br>PPH | Women who<br>had an estimated<br>blood loss of<br>≥500 ml within<br>24 hours after<br>vaginal birth and<br>their partners<br>who were<br>present at the<br>time of the PPH<br>N=11 women, 6<br>partners | Qualitative interview<br>study<br>Maximum variation<br>sampling,<br>explanatory, semi-structured face to<br>face interviews,<br>thematic analysis | Control,<br>communication,<br>consequence<br>(physical and<br>psychological<br>impact),<br>competence |
| Elmir<br>2012a;<br>Elmir<br>2012b;<br>Elmir<br>2012c;<br>Elmir 2014<br>(55-58) | Australia<br>2009 | University<br>campuses and<br>public places such<br>as pharmacies and<br>child care centres | To describe women's<br>experiences of having an<br>EPH following a severe<br>PPH including impacts on<br>early mothering<br>experiences and to explore<br>the way that women find<br>meaning and positivity in<br>their lives | Women who<br>had an EPH<br>following a<br>severe PPH.<br>N=21 | Qualitative interview<br>study using<br>naturalistic inquiry<br>approach<br>Purposive snowball<br>sampling, semi-structured face to<br>face/telephone/e-<br>mail/internet<br>interviews, thematic<br>analysis using<br>inductive method | Between life and<br>death, loss of<br>normality, early<br>mothering<br>experiences,<br>moving forward |
| Knight<br>2016 (39) | United<br>Kingdom<br>2013 | 111 UK hospitals<br>with maternity units | To explore the longer-term<br>health outcomes of women<br>following an EPH to<br>control haemorrhage | Women who<br>had an EPH to<br>control<br>haemorrhage<br>N=78 | Mixed method study<br>using open-ended<br>survey questions<br>Purposive sampling,<br>free text questions at<br>the end of follow-up<br>questionnaire,<br>thematic analysis | The staff<br>providing care,<br>organization /<br>structure of care,<br>longer-term<br>impact of birth<br>experiences |
| Snowdon<br>2012 (61) | United<br>Kingdom<br>2006-2007 | Two hospitals in<br>UK | To explore women's and<br>partners' experiences of<br>severe PPH and its<br>management | Women who<br>had experienced<br>severe PPH and<br>their partners<br>N=9 women, 6<br>partners | Qualitative interview<br>study using<br>interpretative<br>phenomenological<br>approach<br>Purposive sampling,<br>semi-structured face<br>to face interviews,<br>thematic analysis | Confidence vs<br>fear, trust vs<br>mistrust,<br>satisfaction/<br>dissatisfaction,<br>communication<br>difficulties |
| Thompson,<br>2010;<br>Thompson,<br>2011 (12,<br>41) | Australia and<br>New Zealand<br>2006-2007 | 17 hospitals with<br>maternity services<br>in Australia and<br>New Zealand | To identify sources of<br>distress for women and<br>gaps in service provision,<br>particularly their<br>informational needs | Women who<br>had severe PPH<br>N=206 | Mixed-method study<br>using open-ended<br>survey questions<br>Purposive sampling,<br>open-ended self-<br>completed survey<br>questions, thematic<br>analysis | Adequacy of<br>care, emotional<br>responses to the<br>experience,<br>implications for<br>the future, and<br>concerns for<br>their baby |
| Woiski<br>2015 (59) | Netherlands,<br>Not reported | 3 University<br>Hospitals in<br>Netherlands, and<br>childbirth forums<br>on internet | To identify obstacles and<br>facilitators for providing<br>high quality PPH care,<br>from both patient and<br>professional perspectives | Women who<br>gave birth and<br>lost more than<br>1000 ml of<br>blood after birth<br>N=12 | Qualitative interview<br>study<br>Purposive maximum<br>variation sampling,<br>semi-structured face<br>to face/ telephone<br>interviews with<br>patients, framework<br>analysis | Influencing<br>factors from<br>patient<br>perspectives,<br>professional<br>factors and<br>organizational<br>factors |

### Quantitative studies

#### Risk of bias assessment

The results of risk of bias assessment for quantitative studies and quantitative components of mixed-methods studies are presented in **Table 3**. Most included studies (n=15) had a low risk of bias (i.e., low risk in most domains assessed). However, four studies had a high risk of bias in two or more areas of domains assessed, with the main sources of potential bias being participant selection, outcome measurement and incomplete data. (44, 47, 49, 54)

**Table 3.** Risk of bias for quantitative studies using ROBANS (24)

| Study | Participant selection | Exposure measurement | Confounding | Outcome measurement | Incomplete data | Selective reporting |
| --- | --- | --- | --- | --- | --- | --- |
| Chauleur 2008 (8) | + | + | + | + | ? | + |
| Chessman 2018 (32) | + | + | + | + | ? | + |
| Cho 2021 (51) | + | + | + | + | + | + |
| De la Cruz 2016 (44) | − | ? | + | − | + | + |
| Drayton 2016 (33) | + | + | + | − | + | + |
| Eckerdal 2016 (34) | + | + | + | + | ? | + |
| **Eggel 2021;**<br>**Bernasconi 2021**<br>**(54, 60)** | + | + | + | − | − | + |
| Feinberg 2005 (45) | + | + | + | + | − | + |
| Knight 2016 (39) | + | + | + | + | − | + |
| Liu 2021 (42) | + | + | + | + | + | + |
| Michelet 2015 (47) | − | + | − | − | + | + |
| Parry-Smith 2021a;<br>Parry-Smith 2021b<br>(52, 53) | + | - | + | + | + | + |
| Ricbourg 2015<br>(16) | + | + | + | + | - | + |
| Sentilhes 2011<br>(48) | - | + | + | - | + | + |
| Thompson 2010;<br>Thompson<br>2011(12, 40) | + | + | + | + | + | + |
| Thurn 2018 (35) | + | + | + | + | ? | + |
| Ukah 2020 (50) | + | + | + | + | ? | + |
| Van Steijn 2019;<br>Van Steijn 2020<br>(36, 37) | + | + | + | ? | - | + |
| Van Stralen 2018<br>(49) | - | - | - | ? | - | + |
|  | + Low risk of bias , ? Unclear, - High risk of bias |  |  |  |  |  |

#### Results of quantitative synthesis

Quantitative findings are summarised separately for physical and psychological health outcomes for women and their partners in Table 4.

**Table 4.**
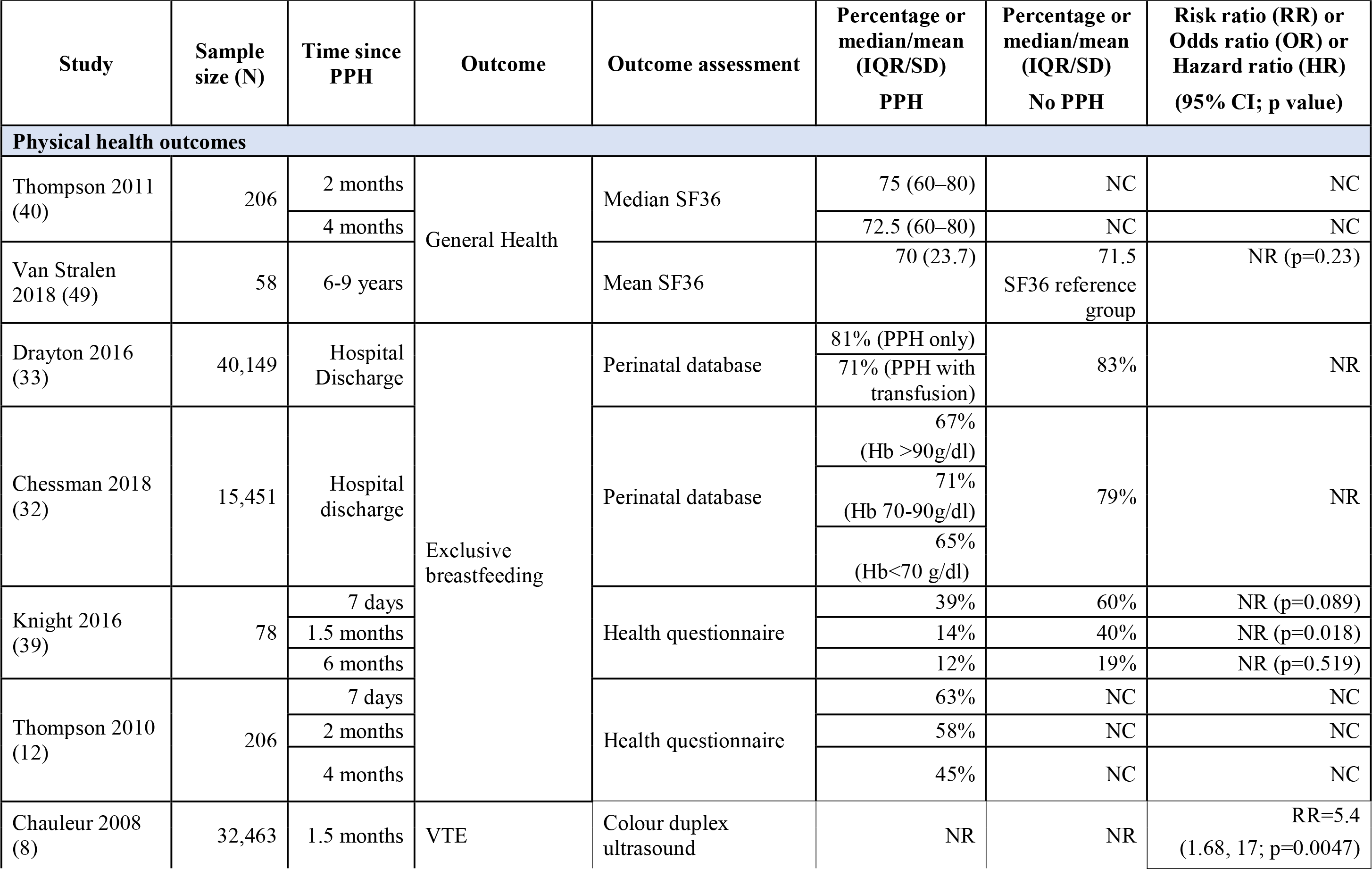
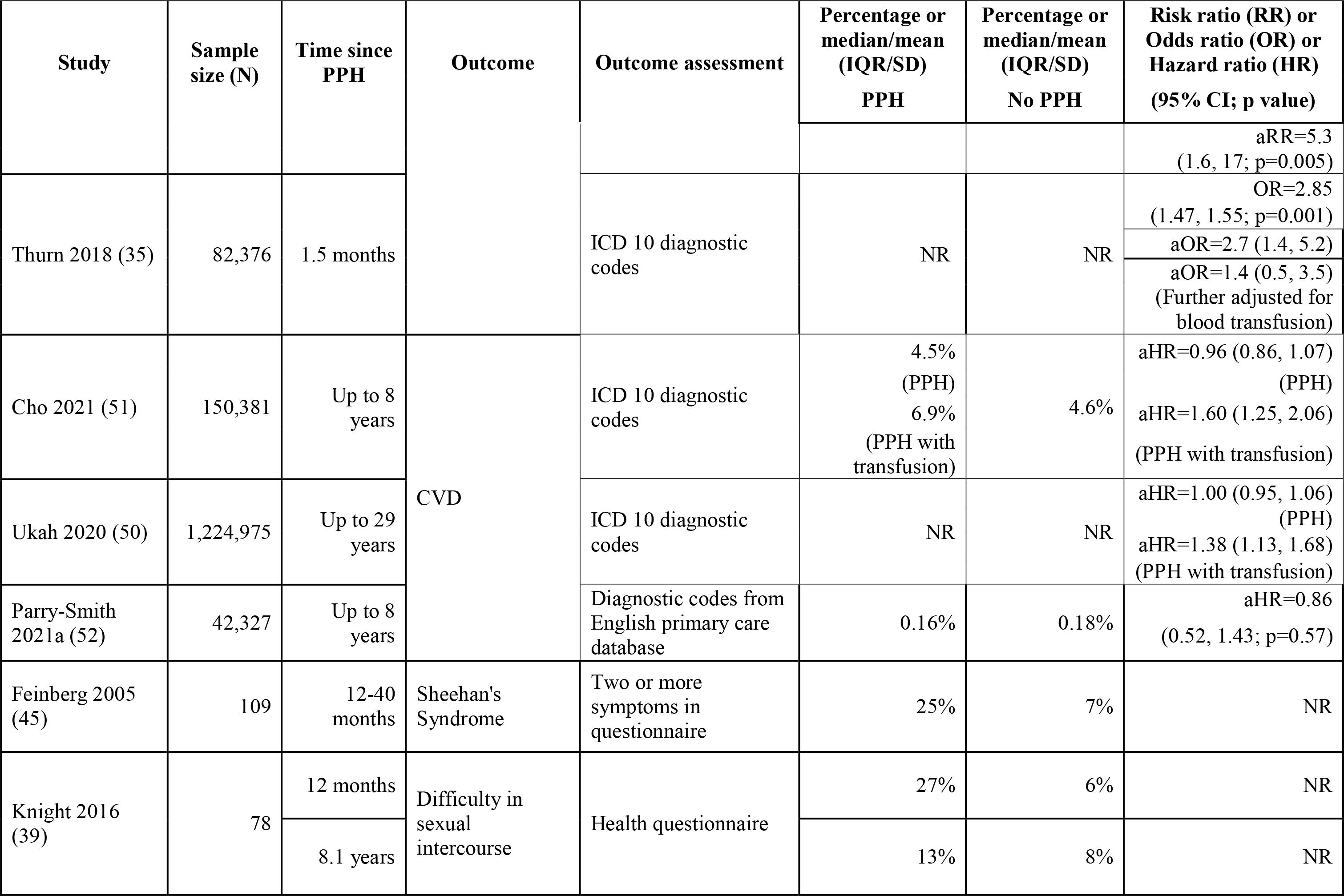
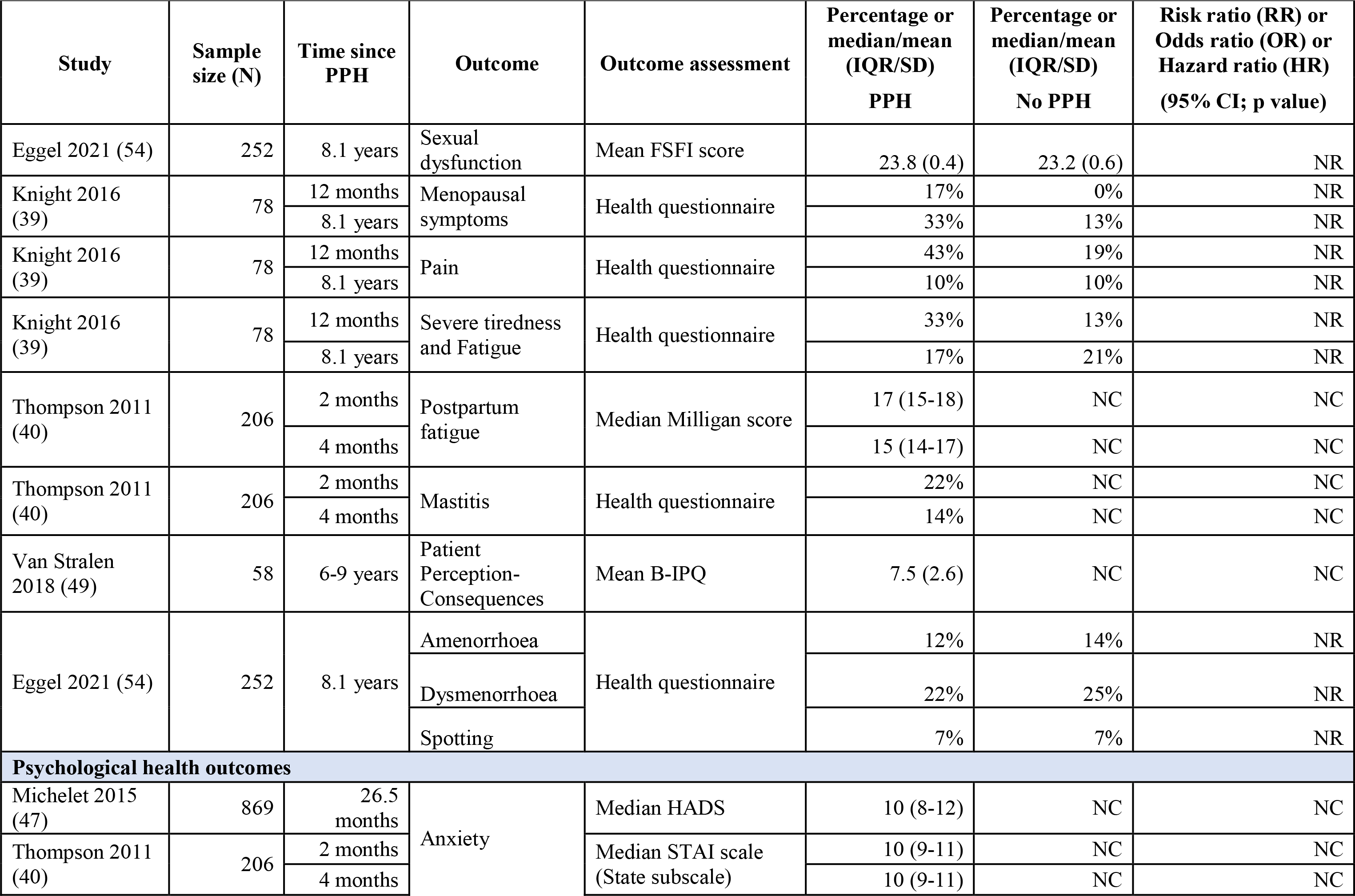
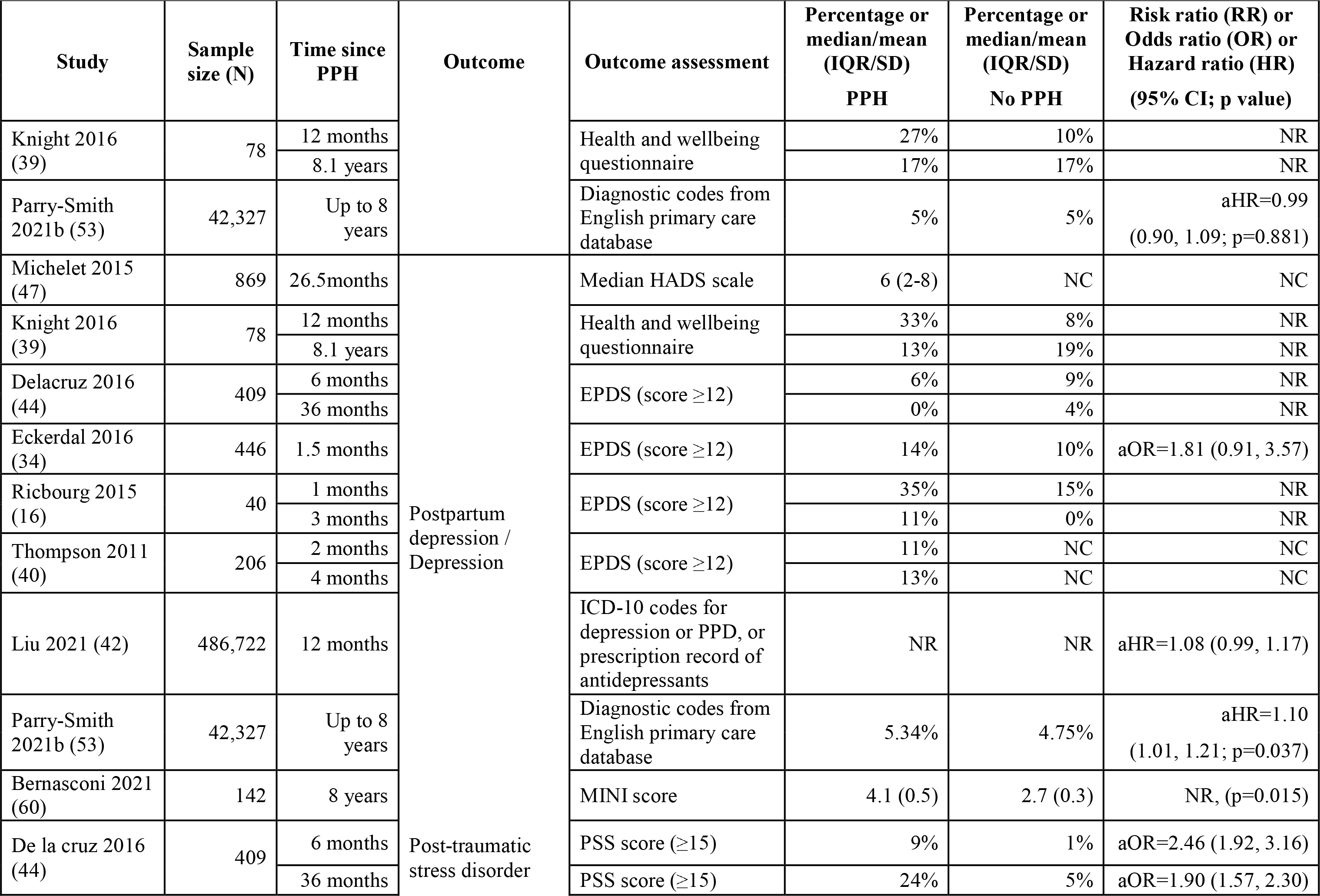
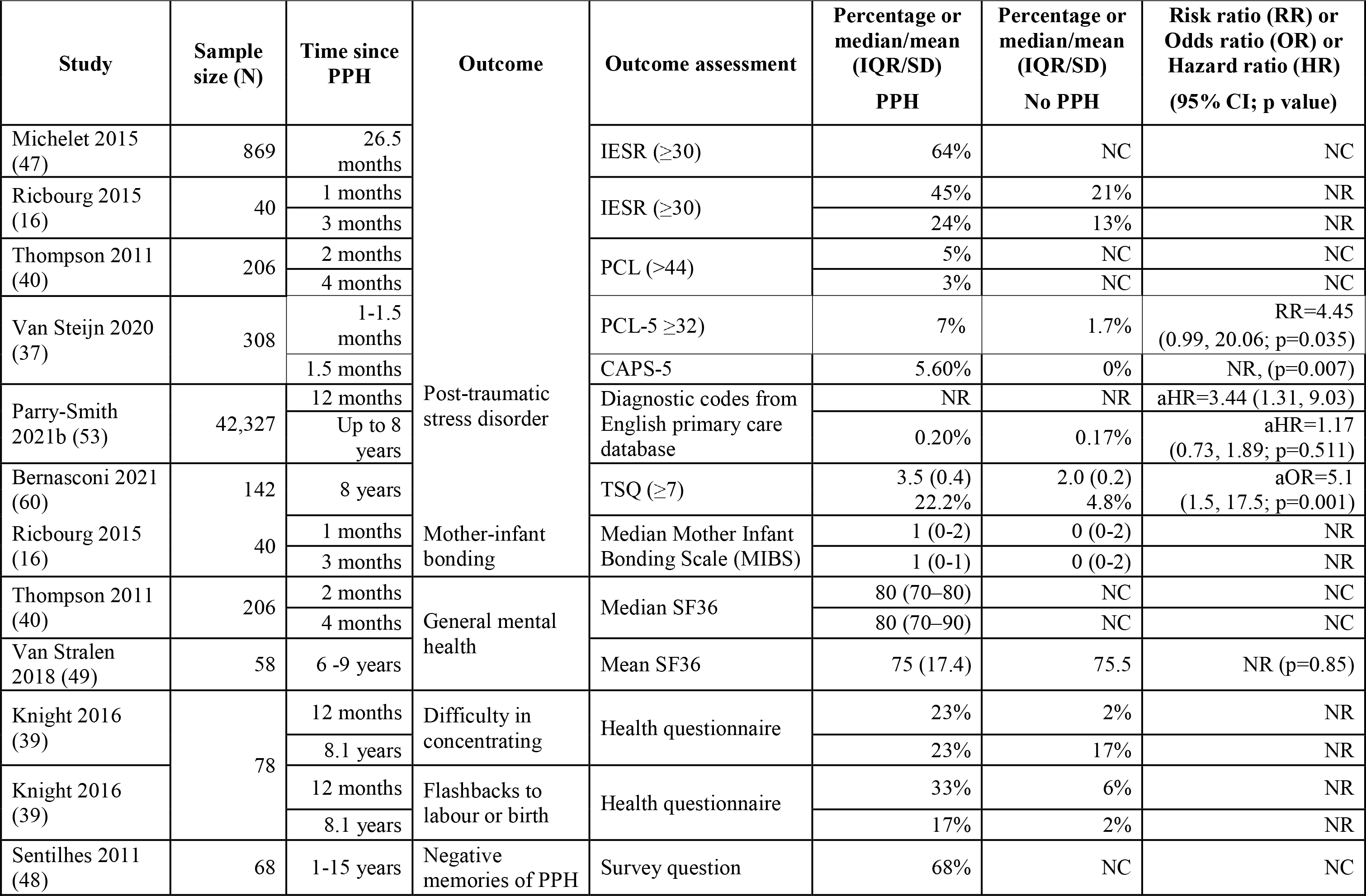
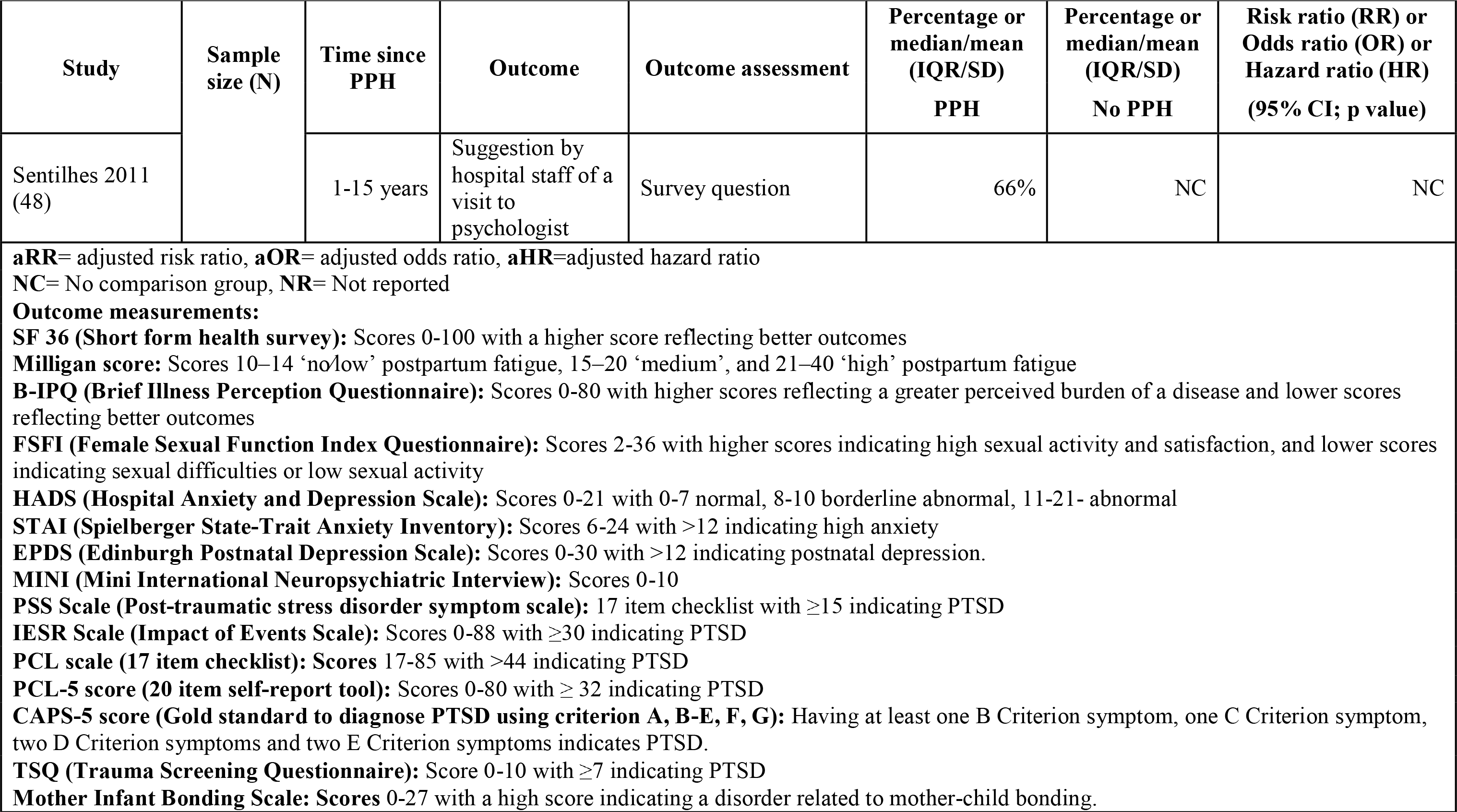
Physical and psychological health outcomes investigated by included quantitative studies

#### Physical health outcomes for women

Two studies reported on general health status in women following PPH. (40, 49) While the first found a decline in general health status at two months and four months following the PPH, (40) the second found that there was no difference in SF36 score for women following PPH compared to the SF36 reference group values. (49) Neither study collected data from an appropriate comparison group.

All four studies that explored breastfeeding as an outcome reported that there was no significant difference in the proportion of exclusive breastfeeding at hospital discharge among mothers who had a PPH, compared with mothers who did not have a PPH. (12, 32, 33, 39) However, the proportion of exclusive breastfeeding at hospital discharge (and at six weeks and six months after birth) was slightly lower among mothers who received a blood transfusion, had a low Haemoglobin level or an EPH following PPH, compared with women who did not have a PPH in three studies. (32, 33, 39) One study did not have a comparison group of women who did not have a PPH. (12)

Two studies explored the potential association between PPH and venous thromboembolism (VTE) at six weeks postpartum. (8, 35) In one study, severe PPH was strongly associated with the occurrence of VTE in the first six weeks after birth after adjusting for caesarean section and low molecular weight heparin use after birth (OR 5.3, 95% CI: 1.6-17, p=0.005). (8) In the second, the authors concluded that PPH was not a major risk factor for VTE unless there was an associated blood transfusion following PPH. (35)

Three studies explored the potential association between PPH and the risk of CVD up to 29 years after birth. (50–52) Two studies suggested that women with PPH requiring blood transfusion had a higher risk of hospitalization for CVD (aHR 1.60, 95% CI 1.25-2.06 and aHR 1.38, 95% CI 1.13, 1.68) compared with women without PPH, after adjusting for potential confounders. (50, 51) However, the third study did not find an association between PPH and hypertension or CVD. (aHR 1.03, 95% CI 0.87-1.22) (52)

Other physical outcomes investigated, each addressed by one study, included Sheehan’s syndrome (damage to the pituitary gland following major haemorrhage), (45) difficulty in sexual intercourse, (39) menopausal symptoms, (39) menstrual disturbances, (54) pain, severe tiredness (39) and fatigue, and mastitis. (12) A higher proportion of women reported these symptoms in severe PPH or EPH groups compared to women who did not have a PPH or EPH.

#### Psychological health outcomes for women

Four studies reported the prevalence of anxiety following PPH at different time points. (39, 40, 47, 53). The exposed group in these studies was women who had a PPH or severe PPH and the follow-up time ranged from two months to eight years after birth. The score for the Hospital Anxiety and Depression Scale (HADS) in the first study indicated moderate anxiety among women who had EPH at median follow-up time of 26.5 months after birth. (47) The second study showed there was mild anxiety among women who had a PPH at both two month and four month follow-up times. (40) Both of these studies did not have comparison groups. The results from the third study indicated women who had hysterectomy were more likely to report anxiety/nerves (27%) compared with women who did not have a PPH (10%) in the first 12 months after birth. Another study, with a follow-up duration of up to 8 years, did not find any association between PPH and anxiety. (53)

Nine studies explored potential associations between PPH and postpartum depression (PPD) or depression. (16, 34, 39, 40, 42, 44, 47, 53, 60) with the follow-up time ranging from one month to eight years after birth. Taken together, the findings of these studies were inconclusive, suggesting different directions of association between PPH and depression. Two studies reported a slightly higher prevalence of depression in the PPH group compared with the non-PPH group at one, three and 12 months after birth. (16, 34) and another two studies with a follow-up of up to 8 years found a higher mean depression score among women who had UAE following PPH, (60) and a 10% increased risk of developing PPD among women who had a PPH compared to women who did not have a PPH after adjusting for age, BMI, smoking, ethnicity, birthweight and delivery method (aHR 1.10, 95% CI 1.01-1.21, p 0.037). (53) The results from multivariable regression models from two further studies suggested no statistically significant association between PPH and PPD at six weeks and 12 months after birth. (34, 42), while a further study reported a slightly higher prevalence of depression in the non-PPH group (6% in PPH vs 9% in non-PPH) at six months after birth. (44) Two studies did not have comparison groups, but one reported a median HAD score of 6 (normal) while the other reported 11% and 13% of women who had a PPH and/or EPH had symptoms of postpartum depression at two and four months after birth. (40, 47)

Seven studies explored the association between PPH and post-traumatic stress disorder (PTSD) at different time points ranging from 1 month to 8 years after birth, indicating a potential association between PPH and PTSD. (16, 37, 40, 44, 47, 53, 60) Five studies found a higher risk of PTSD among women who had a PPH and/or UAE or EPH compared with women who did not experience a PPH. (16, 37, 44, 53, 60) Four of these studies also reported adjusted odds (aOR), risk ratios (aRR) or aHR and these studies suggested a significant association between PPH and PTSD up to eight years after birth [aRR=4.45 (1.5 month); aRR=2.46 (6 months); aHR= 3.44 (within 12 months); aRR=1.90 (36 months); aOR=5.1 (8 years)]. (37, 44, 53, 60) The other two studies did not have comparison groups, but reported respectively 5% prevalence of PTSD among women who had a PPH at two months after birth (40) and 64% prevalence of PTSD among women who had an EPH at the median follow-up of 26.5 months. (47)

Other psychological health outcomes investigated included mother-infant bonding, (16) general mental health score, (40, 49) difficulty in concentrating, flashbacks, (39) negative memories of PPH, and suggestion by hospital staff of a visit to psychologist. (48) Results indicated that a higher proportion of women in severe PPH or EPH groups reported having difficulties in concentrations and flashbacks compared with women who did not have a PPH or an EPH.

#### Physical and psychological health outcomes for partners

Three quantitative studies reported on the prevalence of psychological health outcomes among partners of women who had experienced PPH. (36, 49, 60) The first study reported that there was no strong association between PTSD diagnosis and witnessing PPH among partners of women according to a self-report questionnaire. (36) The second study explored the quality of life for partners of women who had experienced severe PPH using SF-36 general health survey tools. (49) The results indicated that partners of women who had severe PPH had better scores overall compared with the reference group’s scores from myocardial infarct and systemic lupus erythematosus patients. In contrast to these studies, the third study reported that there was a higher prevalence of depression and PTSD among partners of women who had PPH compared to partners of women without PPH (11.5% versus 1.5%, p=0.019). (60)

### Qualitative studies

#### Quality assessment

The results of quality assessment for the eight included qualitative studies and qualitative components of mixed-methods studies are presented in **Table 5**. All but one of the studies had a response of “yes” to at least seven of the questions in the CASP tool, with the remaining study, having a “partially” response for three questions. (39) Only two of eight studies (46, 61) reported the relationship between researchers and participants clearly (i.e., Q6), so it was not possible to assess whether the roles of researchers had a potential impact on the research study and interpretation of findings.

**Table 5.**
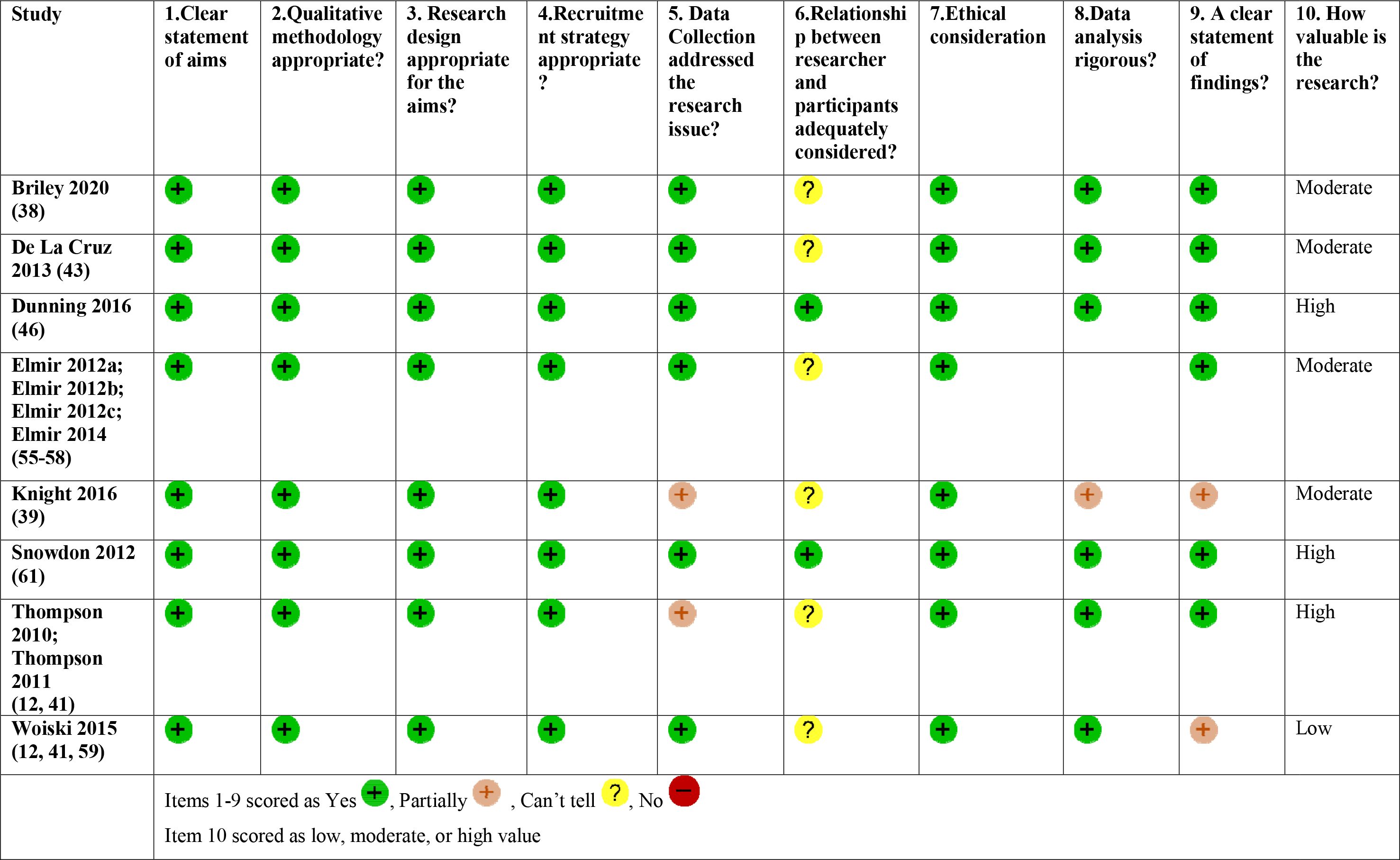
Quality assessment for qualitative studies using CASP tool (25)

#### Results of qualitative synthesis

Analytical themes are presented under four headings: physical impact, psychological impact, psychosocial impact, and experiences of care. **Fig 2** presents the overall structure of the themes arising from the synthesis, organized into four key time periods: during the emergency, immediate hospital recovery, postnatal period up to six months after birth, and longer-term.

**Fig 2.**
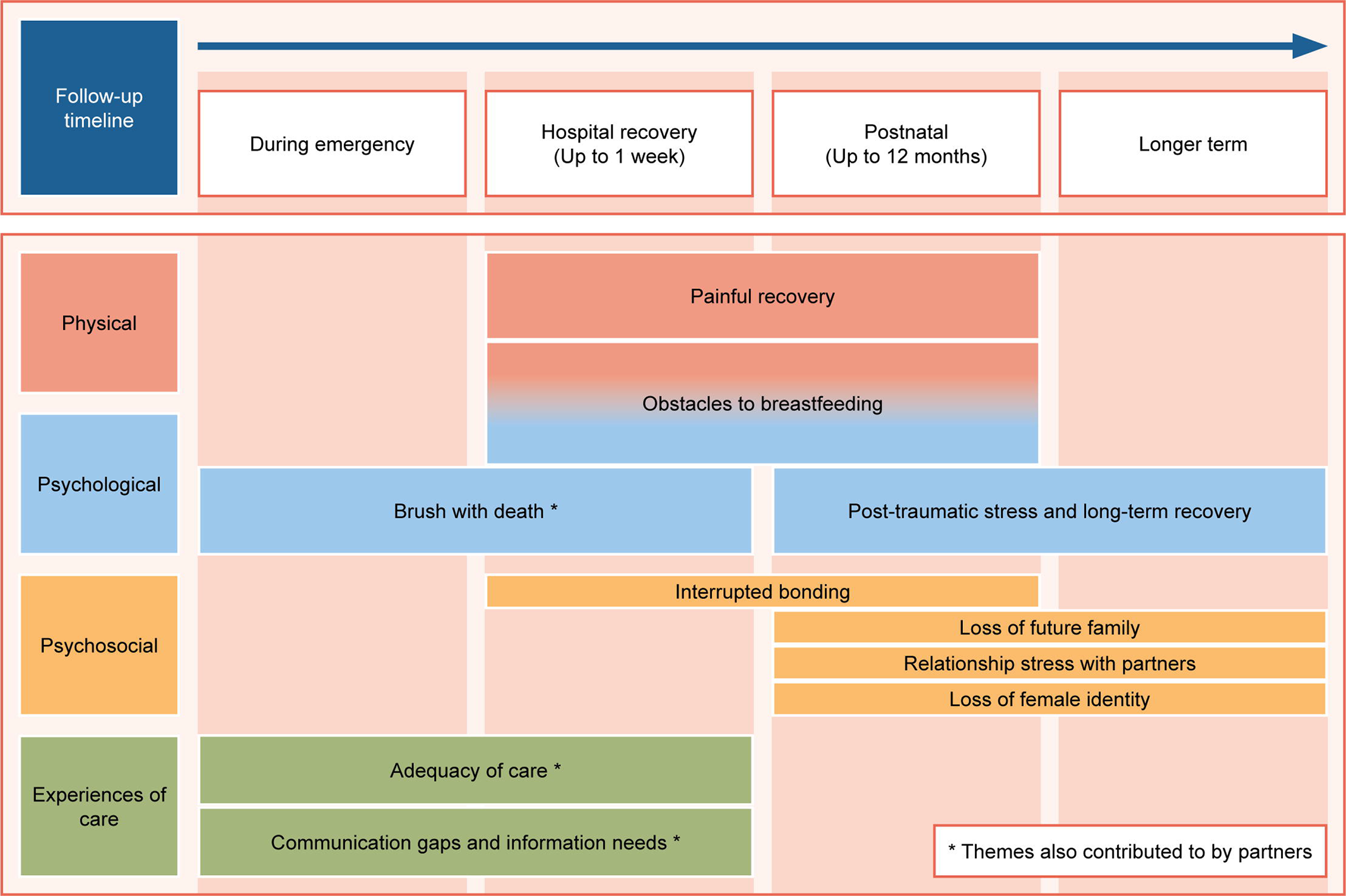
Summary of analytic themes from thematic synthesis of qualitative studies (n=8)

### Women’s experiences

#### Physical impact

##### Painful recovery

This theme was contributed to by five studies and encompasses women’s experiences of pain at the time of the PPH and for up to a year afterwards. (39, 41, 43, 46, 56) Physical limitations during the immediate postnatal period and beyond were consistently reported by women across five studies. (39, 41, 43, 46, 56) Their descriptions indicated that pain prevented them from performing simple day to day activities such as walking and cooking dinner. (46, 56) One particular concern raised by women was that they were not able to take proper care of their babies as they were not fully recovered from pain. (41) In the longer-term, one woman who experienced an EPH reported that it took a year for her to physically recover. (39)

##### Obstacles to breastfeeding

This theme was contributed to by three studies. (12, 43, 55) During their immediate hospital recovery, women reflected that their initial separation from their baby when they were admitted to intensive care was one of the key barriers to initiating breastfeeding. (12, 55) Other key obstacles reported by women included pain, fatigue, limited mobility and posture problems, and delayed milk production. (12, 55) Women also talked about how their efforts and willingness to breastfeed after PPH were not sufficiently supported by health care professionals, including for example being told that they should not be bothered to breastfeed after a traumatic childbirth (12, 43)

#### Psychological impact

##### Brush with death

This theme was contributed to by three studies and encompasses women’s acute psychological distress as a result of an unexpected encounter with an emerging life-threatening event. (41, 43, 56)

For many women, being so close to death was traumatic and frightening, particularly when women did not anticipate that death was a possibility. (41, 43, 56) Their, overwhelmingly negative, emotional responses to this were described as varying from “shock”, to having a “major anxiety attack” or having an “out of body experience”. (41, 43, 56) Elmir et al. (2012) also highlighted that women were afraid of “leaving their young children behind to the unknown” and not knowing “whether or not they would cope without a mother”.

##### Symptoms of post-traumatic stress and long term recovery

This theme was contributed to by five studies and describes women’s experiences of post-traumatic stress symptoms (PTSD) following PPH. (39, 41, 43, 46, 56, 62) PTSD is characterised by four key symptoms: reliving experiences, avoidance of reminders of trauma, negative thoughts and mood, and hyperarousal. (14)

Women described re-experiencing their traumatic birth in the form of vivid memories, flashbacks and nightmares. (41, 56) Triggers for flashbacks included seeing other pregnant women, being in a hospital environment or seeing stories in the media about giving birth. (56)

Women also talked about avoiding reminders of trauma. One woman, for example, said that she avoided hospital environments. (56) For some women, reported in two studies, these symptoms were severe, including having suicidal thoughts. (39, 56)

While the overwhelming majority of women’s observations from most studies were negative, women from one study reflected on being able to take something positive from their experiences, talking about how it helped them find some perspectives and resilience. (62)

#### Psychosocial impact

##### Interrupted bonding

This theme was contributed to by three studies and reflects women’s perceptions of bonding with their infants and their role as a mother during their recovery in hospital and in the postnatal period. (12, 43, 55) This theme is interlinked with the theme: obstacles to breastfeeding.

Women reported feeling that bonding with their babies was disrupted by physical separation because of their admission to intensive care or their babies’ admission to neonatal care. Women talked about having concerns for their babies’ wellbeing and not knowing if they were okay. (12, 55) Women reflected that this initial separation prevented them from holding, touching and breastfeeding their babies, and many felt that this was a key barrier establishing a close relationship with their babies.

Women’s reports in these studies showed that this initial interrupted bonding could continue in the weeks following hospital discharge for mothers, some of whom experienced extreme tiredness, continuing for up to a year after childbirth. Some women in one study talked about feeling “upset” and “guilty” when their caring responsibilities were taken away by their family members and partners. (55)

##### Loss of future family

This theme was contributed by two studies and encompasses women’s feelings of loss in relation to future childbearing following PPH. (39, 41) At four months after childbirth, some women talked about feeling worried about having a PPH in any future births and losing their confidence in their ability to go through birth again. (41) Traumatic birth experiences convinced some women not to have more children. (41) Women who had a hysterectomy following PPH talked about their regret at not being able to have more children. (39)

##### Relationship stress with their partners

This theme was contributed to by three studies and reflects women’s perceptions of their relationships with their partner following PPH and/or hysterectomy, in the immediate postnatal period and up to a year after birth. (39, 46, 57) In one study, one woman who had a PPH described having a relationship breakdown with her partner following a traumatic birth. (46) Women who had a hysterectomy following PPH talked about sexual tension and intimacy issues with their partners. (39, 57) For instance, a woman in one study mentioned loss of libido and avoidance of physical contact with her partner, (39) with this lack of sexual desire and “fear for intimacy” also being echoed by women in a second study. Women also reported developing insecurities over time causing a strain in their marriage. (57)

##### Loss of female identity

This theme was contributed to by two studies and covers women’s experiences of hysterectomy as a consequence of a PPH. (39, 57) Women’s accounts reflected that the memories lingered with them for many years and this changed how they felt about themselves as a woman. (57) For some women the lack of uterus was associated with feelings of incompleteness and emptiness. (57) This was echoed by women in the second study as they talked about how hysterectomy made them feel like a different person and not like a “complete woman”. (39)

#### Experiences of care

##### Adequacy of care

This theme was contributed to by six studies. Both positive and negative experiences of care were reported by women, but all demonstrated the crucial role of health care professionals helping women feel calm and supported in an emergency when they established mutual trust. (38, 39, 41, 43, 46, 59) Some women reflected that they appreciated it when the doctors and midwives appeared confident and calm, and took time to reassure them afterwards. (38, 46) Others described feeling unsupported when midwives were “impatient” towards them, “too busy” to explain things or did not “acknowledge” them while providing care. Women’s accounts also showed how the care environment could affect their overall experiences. One woman, for example, reported that she was very pleased to be in the critical care unit compared to a busy ward. (39)

Insufficient follow-up care and limited resources for psychological health during the postnatal period were identified in four studies. (38, 39, 41, 43) Women’s accounts indicated a need for psychological counselling and follow-up debrief during the postnatal period. (39, 41, 43) Some women also mentioned that patient information websites and support for breastfeeding would be helpful for them. (41, 59)

##### Communication gaps and information needs

This theme was contributed to by six studies and describes women’s information and communication needs and preferences, including their information seeking behaviours. (38, 39, 41, 43, 46, 59, 61) It is important to provide women who had experienced PPH a thorough explanation and clear communication about their condition, treatments received including hysterectomy and its potential consequences. (39) A lack of information about their current health condition could worsen psychological distress for women and their partners. (43, 61)

During the emergency, women appreciated receiving a full explanation and information about their hysterectomy and the indications for it before they consented to have an EPH. Women from five studies consistently reported that they would have liked to know about available treatment options and consequences of PPH, including recovery time after they had experienced PPH. (38, 41, 43, 46, 61)

During the immediate recovery period, women’s accounts demonstrated that most women were unaware of excessive blood loss. (38, 46) They wanted information about what had happened to them, their current location (e.g. ICU or postnatal ward), and the condition of their babies in simple terms without using confusing technical words. (41, 43, 61) Another communication gap reported by women who had a hysterectomy was that nurses were not aware of their complicated birth and women found it “unsettling” when they had to explain to that they did not have a uterus for examination, and had no need for contraception. (43)

Women’s information needs extended into the postnatal period, and included communication about the implications of PPH and recovery time, impacts on breastfeeding, and available support services. (41, 43, 46, 59)

#### Partners’ experiences

Three qualitative studies also included partners of women as participants. (38, 46, 61) Partners’ accounts showed that, while women were facing life and death situations, partners also experienced psychological distress from witnessing this. (46, 61) One partner described himself as “desperate” at being left alone with his baby without support or communication about what had happened to the baby’s mother. (61) This was also echoed by others as “distressing”, and one partner reported seeking help from the GP and counselling services in the months after witnessing such a traumatic birth. (46)

During the emergency and the immediate recovery period, partners appreciated it when doctors took time to reassure them and did not find it helpful when health care staff appeared to show panic or went quiet. (38) In terms of information and communication needs and preferences around the time of the PPH, (38, 46, 61) partners wanted more information about available treatment options and potential implications while waiting to find out what had happened. (46, 61) They reported feeling frustrated when they were being asked to leave the emergency room without being fully informed about what was happening. (46, 61) Some suggested that an information sheet about PPH would be helpful for partners and family members to be able to learn more about the situation. (61)

## Discussion

The main aim of this review was to identify and summarise the available quantitative and qualitative evidence about longer-term physical, psychological and psychosocial health outcomes for women and their partners following primary PPH in high-income countries. We included 24 studies with a mix of methodological quality ranging from weak to strong. Follow-up time ranged from hospital discharge following PPH to 29 years. Among these, nine studies explored the longer-term impacts of primary PPH after five years, and just two quantitative studies (48, 50) and one qualitative study (55–58) had a follow-up period longer than ten years. Therefore, there is limited research about longer-term health outcomes following primary PPH compared with the many studies that have investigated acute morbidity and health outcomes related to primary PPH. (7)

Our review provides evidence that women with PPH are more likely to have persistent physical and psychological health problems during the postpartum period compared with women who have not had a PPH. These differences were more pronounced for women who had some indication of severe PPH such as a blood transfusion, or an EPH. The quantitative findings suggest that women with severe PPH are more likely to have a long recovery time, low general health score, symptoms of tiredness/fatigue, and are less likely to exclusively breastfeed. These quantitative findings are consistent with women’s descriptions of their physical functioning and limitations in performing simple day-to-day activities and women’s experiences of challenges around initiating and maintaining breastfeeding from qualitative studies. Our review’s findings build upon the findings of a previous review conducted in 2016 by including updated literature and by synthesizing available qualitative evidence on women’s and partners’ experiences of PPH illuminating the different directions and size of effects in quantitative studies. (14)

A survey conducted among 372 women who had major obstetric haemorrhage in the Netherlands, in which PPH was not distinguished from an antepartum haemorrhage, indicated that 28% of the participants experienced longer-term negative impacts up to six years after birth. (63) Major obstetric haemorrhage was found to have a negative impact on the partner and family, and on their work, and 25% of these women reported having an additional absence from work in addition to maternity leave. (63)

Bearing in mind the limitations identified in terms of the quantity and quality of the evidence, our review indicates that some longer-term physical and psychological health outcomes after PPH may be severe and extended several years after childbirth. Included quantitative studies indicated that the prevalence of PTSD symptoms ranged from 3% to 64% among women with PPH. Women’s descriptions from qualitative studies, which included key symptoms of PTSD such as flashbacks and nightmares of having PPH, were supportive of these quantitative findings. These PTSD symptoms were commonly present among women who had severe PPH and lasted for years after birth. This finding is supported by qualitative studies which concluded that childbirth related PTSD can have severe and lasting effects on women and their relationships with family. (64–66) In the general population, the mean prevalence of postpartum PTSD is 4%-6% and may be higher, with a mean prevalence of 19% among women with physical complications and mental health problems such as depression. (67, 68) While our review’s findings are indicative of a higher prevalence of PTSD symptoms following primary PPH, (16, 37, 44, 60) more evidence is required before concluding a causal association between PPH and PTSD. (18)

Our review included three studies which explored the association between PPH and CVD, and the findings suggested that women who had a severe PPH requiring a transfusion were more likely to develop CVD in later life. Prior studies have suggested that women with severe PPH are at a higher risk of acute heart failure and myocardial ischaemia which results from haemorrhagic shock. (69, 70) Therefore, there is a potential that this acute failure has implications for the cardiovascular system which may put women at higher risk of developing subsequent CVD in later life. Other obstetric complications, including pre-eclampsia, preterm birth, maternal anaemia, and gestational diabetes, have also been shown to increase the subsequent risk of developing CVD. (71–74) It is therefore important to consider women’s obstetric history before reaching conclusions about a casual association between PPH and CVD.

Qualitative evidence included in this review indicated that whether women received adequate support during the immediate recovery and postnatal period may affect their acute psychological wellbeing and have an impact on longer-term psychological health outcomes after PPH. For example, women’s accounts suggested that they received insufficient follow-up care and had unmet needs for breastfeeding and psychological health services. Health care professionals and family members were crucial in helping women feel calm and supported. This is supported by qualitative studies conducted in Australia which have described how care providers’ interactions and postnatal support from partners can affect women’s experiences of trauma during childbirth. (75–77)

We found very limited evidence about outcomes for partners after PPH, with only three quantitative studies without comparison groups. The evidence from one study suggested an increased risk of PTSD among partners who witnessed PPH while another two studies did not indicate any strong associations between PTSD or general health status. However, partners’ accounts in qualitative studies indicated that they experienced acute psychological stress and anxiety due to inadequate information when women were in the emergency room, and some of them required counselling services after witnessing PPH. PPH may also have an impact on relationships between women and their partners. Witnessing their partner having PPH and having relationship difficulties may affect partner’s psychological wellbeing beyond the postnatal period. Qualitative studies into fathers’ experiences who witnessed birth trauma have identified that this can affect their mental health and relationships with friends and family long into the postnatal period. (78, 79) Studies have also found that men are more likely to cope with these traumatic experiences by avoidance, and may be reluctant to seek necessary care which may lead to poor psychological health outcomes. (80, 81) Therefore, it is crucial to investigate the potential psychological impact among partners who have witnessed a PPH.

### Strengths and limitations

To our knowledge this is the first mixed-methods systematic review integrating quantitative and qualitative evidence to explore the potential longer-term health outcomes of primary PPH. The inclusion of qualitative evidence about women’s and partners’ experiences helped illuminate our understanding of findings from the quantitative studies. A comprehensive and reproducible search strategy was conducted, the methods were rigorous, and quality assessment and data extraction were performed by two authors independently of each other.

Many of the limitations of this review reflect shortcomings of the included studies. First, there were some challenges associated with drawing conclusions about the association between PPH and some health outcomes because most included studies did not have appropriate comparison groups and conclusions were made from the prevalence/proportions among the exposed group without consideration of potential confounders. For most studies, the follow-up time was less than five years. Most studies used self-reported health questionnaires, rather than objective measurements, such as clinical diagnosis.

At the review level, the heterogeneity of studies, including different definitions of PPH, different follow-up time and variation in the outcomes investigated, meant that it was not possible to conduct meta-analysis. Additionally, the number of studies that made an attempt to establish the potential longer-term impact of PPH was limited. Due to resource limitations, we were unable to consider for inclusion five potentially eligible papers that were not written in English.

### Implications for research

There is limited evidence about the longer-term impact of primary PPH on women and their partners, hence there is a need for further research in this area. In particular, cohort studies with comparison groups, and longer duration of follow-up should be conducted to explore the association between PPH and selected chronic health outcomes, for instance, CVD. Study designs could be improved by using standardised outcome assessment such as applying standardised self-report tools validated for this population or clinical interviews. Additionally, emphasis on the potential cumulative impact of PPH throughout the obstetric history of women should be considered. This is particularly important in multiparous women who may have more than one primary PPH with a potentially more severe impact. As evidenced by qualitative studies, differences in quality of health care (e.g., frequency of follow up, access to counselling and psychologist) should be also considered in any future prospective cohort studies.

In spite of increasing trends in PPH, women are less likely to experience adverse fatal outcomes due to better management and recording of severe PPH compared to the past decade. (4, 82) Globally, the research focus on other near-miss complications has shifted from maternal mortality to surveillance of the burden of severe maternal morbidity, including improvement in the quality of follow-up care. (83, 84) The same principle should be applied to future research on PPH to explore whether the trajectory of PPH and its acute morbidities is associated with subsequent risks of developing chronic disease.

More research should also be conducted to explore the potential longer-term psychological implications for partners witnessing PPH. Qualitative evidence in this review suggested signs of acute distress, with significant information needs and follow up support needs for partners who witnessed women experiencing PPH. Obtaining information about partners’ mental health and coping strategies could be beneficial to both the woman and her partner in terms of identifying support needs moving forward.

## Conclusions

This review synthesized evidence about longer-term physical and psychological health outcomes among women who had a primary PPH in high-income countries, and their partners. While the available evidence mainly focuses on immediate health outcomes following birth, this review suggests that women can have longer-term health problems associated with PPH beyond one-year after childbirth. The extent of the impact of these health outcomes is poorly researched and may be influenced by the severity of PPH, presence of other obstetric complications, and the quality of care received. The limitations of the evidence about longer-term health outcomes after PPH emphasizes the need for further research in this area.

## Supporting information

S1 Checklist

## Data Availability

All relevant data are within the manuscript and its Supporting Information files.

## Acknowledgements

We would like to express our sincere appreciation to Nia Roberts (Senior librarian, University of Oxford Bodleian Health Care Libraries) for her guidance in developing search strategies.

## Supporting Information

**S1 Checklist. PRISMA 2020 checklist.**

**S1 File. Medline Search Strategy.**

**S2 File. Data extraction forms**

**S1 Table. Modified risk of bias assessment tool for non-randomized studies (ROBANS)**

