## Supplementary material for "Primary postpartum haemorrhage and longer-term physical, psychological, and psychosocial health outcomes for women and their partners: a mixed-methods systematic review": S1 File

**S1 File. Search strategy for Medline (Ovid)**

| # | Searches |
| --- | --- |
| 1 | exp Postpartum Hemorrhage/ |
| 2 | ((postnatal or post-natal) adj2 h?emorrhag*).mp. |
| 3 | ((peripart* or peri-part*) adj2 h?emorrhag*).mp. |
| 4 | ((postpart* or post-part*) adj2 h?emorrhag*).mp. |
| 5 | ((postnatal or post-natal) adj2 (bleeding or blood loss)).mp. |
| 6 | ((peripart* or peri-part*) adj2 (bleeding or blood loss)).mp. |
| 7 | ((postpart* or post-part*) adj2 (bleeding or blood loss)).mp. |
| 8 | (obstetric adj2 h?emorrhag*).mp. |
| 9 | (obstetric adj2 (bleeding or blood loss)).mp. |
| 10 | ((postnatal or post-natal) adj5 transfus*).mp. |
| 11 | ((peripart* or peri-part*) adj5 transfus*).mp. |
| 12 | ((postpart* or post-part*) adj5 transfus*).mp. |
| 13 | ((postnatal or post-natal) adj10 hysterectom*).mp. |
| 14 | ((peripart* or peri-part*) adj10 hysterectom*).mp. |
| 15 | ((postpart* or post-part*) adj10 hysterectom*).mp. |
| 16 | ((postnatal or post-natal) adj10 morbidit*).mp. |
| 17 | ((peripart* or peri-part*) adj10 morbidit*).mp. |
| 18 | ((postpart* or post-part*) adj10 morbidit*).mp. |
| 19 | 1 or 2 or 3 or 4 or 5 or 6 or 7 or 8 or 9 or 10 or 11 or 12 or 13 or 14 or 15 or 16 or 17 or 18 |
| 20 | (("semi-structured" or semistructured or unstructured or informal or "in-depth" or indepth or "face-to-face" or structured or guide) adj2 (interview* or discussion* or questionnaire*)).tw,kw. |
| 21 | (focus group* or qualitative or ethnograph* or fieldwork or "field work" or "key informant").tw,kw. |
| 22 | interviews as topic/ or focus groups/ or narration/ or qualitative research/ |
| 23 | 20 or 21 or 22 |
| 24 | epidemiologic studies/ |
| 25 | (observational adj (study or studies)).tw. |
| 26 | exp case control studies/ |
| 27 | exp cohort studies/ |
| 28 | Case control.tw. |
| 29 | (cohort adj (study or studies)).tw. |
| 30 | Cohort analy$.tw. |
| 31 | (Follow up adj (study or studies)).tw. |
| 32 | Longitudinal.tw. |
| 33 | Retrospective.tw. |
| 34 | Cross sectional.tw. |
| 35 | Cross-sectional studies/ |
| 36 | (case$ and series).tw. |
| 37 | 24 or 25 or 26 or 27 or 28 or 29 or 30 or 31 or 32 or 33 or 34 or 35 or 36 |
| 38 | 23 or 37 |
| 39 | 19 and 38 |
