## Supplementary material for "Primary postpartum haemorrhage and longer-term physical, psychological, and psychosocial health outcomes for women and their partners: a mixed-methods systematic review": S1 Table

**S1 Table. Modified risk of bias assessment tool for non-randomized studies (ROBANS)**

|  | Domain | Adapted criteria | |
| --- | --- | --- | --- |
|  |  | Low risk of bias^1^ | High risk of bias^1^ |
| 1 | Participant selection | The following criteria should be considered:  **Cohort studies**   - Women with and without PPH were selected from the same population group(s) and time period(s) (appropriate comparison group) - Cohort studies: The cohort started with healthy (i.e. outcome free) participants and assured that people with the outcomes occurring before pregnancy / at the time of birth were excluded.   **Case-control studies**   - Only incident cases were included (e.g. newly diagnosed/treated cases) - The selection of cases and controls was independent of the exposure status (i.e. PPH), and cases and controls were derived from the similar source population, and the source of controls was population-based. - Observational study designs with a proper comparison group (e.g. women with or without PPH were followed up over time; exposures were assessed for cases and comparable controls)   **Cross-sectional studies**   - Selection/participation was independent of the exposure (i.e. PPH) and outcome status | **Cohort studies**   - Women with or without PPH were selected from different population groups and/or time periods (e.g. historical unexposed cohort) - Participants had the outcomes at baseline or they were likely to have undiagnosed outcomes.   **Case-control studies**   - Both the incident and prevalent (pre-existing) cases were included. - The selection of cases and control was dependent of the exposure status, or the cases and controls were not selected from the similar source population, or the source of controls was not population-based. - Observational study designs without a proper control group (e.g. case series or exposed only cohorts)   **Cross-sectional studies**   - A clear case of selection bias. Selection / participation varied by a combination of exposure and outcome (e.g. women with PPH who were mentally unfit did not participate in the survey) |
| 2 | Confounding variables | The major confounding variables were adequately considered and adjusted in the study design (through participant restriction/exclusion or matching) and/or in the analysis. Such confounders might include, for example   1. Maternal age at the time of birth 2. Maternal education or socioeconomic status (or family socioeconomic status) 3. Maternal BMI (at first antenatal visit) or gestational weight gain 4. Parity 5. Maternal smoking habit   Other major confounders include:   6. Presence of other obstetric complications (e.g. pre-eclampsia, gestational hypertension/diabetes, prolonged third stage of labour) 7. Neonatal condition (e.g. admission to neonatal unit, baby weight or outcomes of pregnancy) | The major confounding variables were not considered in the study design or analysis (i.e. the study did not confirm or adjust for the potential confounders such as maternal age, maternal education or socioeconomic status, BMI, Parity etc.) |
| 3 | Exposure measurement | The following criteria should be considered:   - PPH was defined using standardised diagnostic criteria, for example using definitions of WHO, RCOG, ACOG, or ICD-10 codes. - The amount of blood loss was assessed prospectively and in a standardised manner by a qualified/trained health care professional. | Any of the following:   - PPH was defined using non-standard or unclear criteria. - The measurement of PPH was not comparable among all study participants. - The data on PPH was self-reported.   For case-control/cross-sectional studies, please consider:   - A clear case of interviewer bias (e.g. The interviewer knew the case status and the exposure of interest (PPH), so they asked questions differently to women with outcomes) - A clear case of recall bias (e.g. Participants were asked the history of PPH 10 years after birth and cases (women who are unwell) recalled PPH more frequently than controls.) |
| 4 | Blinding and outcome measurement | The following criteria should be considered:   - The outcome was evaluated using established diagnostic/classification criteria or validated tool (e.g. Post-natal depression scale) or laboratory tests - Outcome evaluation was performed in a standardised fashion by qualified assessors (e.g. midwives, doctors, psychologists) - The duration of the follow-up is reasonably long enough for the outcomes assessed to occur. - The timing of outcome assessment was comparable for all participants - Outcome assessors were blinded to women’s medical history/record for PPH or were blinded to the research question. If blinding is absent, it is less likely to be the source of differential measurement error (e.g. the outcome measurement was based on the laboratory test, or existing records) | - The outcome was defined using non-established (e.g. ad hoc) diagnostic criteria/tools with uncertain validity/reliability (including self-reporting using such tools) or diagnostic criteria were not clear - In cohort studies, the outcome was evaluated retrospectively from medical records that were not validated or did not have quality checks. - The outcome was assessed using procedures or diagnostic criteria, which were not comparable for all the study participants - The duration of outcome assessment was not long enough for outcomes to occur. - The timing for outcome measurement was not comparable for all participants - Evaluation of the outcome was unblinded to the women’s medical history when the absence of blinding can be the source of potential measurement errors |
| 5 | Incomplete data | The following criteria should be considered:   - The proportion and causes of missing data were similar between exposed and unexposed groups (women with or without PPH), or between cases and controls - In cohort studies, women lost to follow-up were comparable to women that were not lost to follow-up regarding characteristics relevant to the study outcome (i.e. non-differential LTFU) - The potential impact of missing data was corrected in the analysis - The study results were demonstrated to be robust to the presence of missing data (e.g. via sensitivity analyses) | - Women lost to follow-up and those who were not lost to follow-up differed in characteristics deemed relevant to the study outcome (differential LTFU) (e.g. LTFU was more common in women with PPH because they were emotionally unfit) , and these differences were not addressed in the analysis. - The proportion and causes of missing data differed between participants with or without PPH (or cases and controls) , and these differences were not addressed in the analysis - The study results were influenced by the missing data |
| 6 | Selective reporting  (within study) | The following criteria should be considered   - If a study protocol is available, the pre-defined primary/secondary analyses were described as planned or justified in the study report if there were any legitimate changes from the protocol. - Analyses of all of the outcomes/exposures that are relevant to the study objectives were reported (e.g. check the outcomes mentioned in the methods section and the reported results, or consider missing report of the measures which are usually collected together e.g. SBP and DBP) - The effect estimates reported do not seem to have been selected from (i) alternative outcome/exposure measurements, (ii) multiple analytic approaches, or (iii) multiple subgroups | - The pre-defined primary analyses were not fully reported or reported as planned in a protocol or not justified for the differences. - Additional analyses that were not pre-specified or directly relevant to the study objectives were included (e.g. subgroup analyses, secondary variables), except for those with a clear justification - Some analyses of exposures/outcomes that are relevant to the study objectives were not reported - The effect estimates reported seem to have been selected from (i) alternative outcome/exposure measurements, (ii) multiple analytic approaches, or (iii) multiple subgroups |

^1^ If there is insufficient information to determine whether the risk of bias is low or high, it should be classified as “unclear”
