## Supplementary material for "Primary postpartum haemorrhage and longer-term physical, psychological, and psychosocial health outcomes for women and their partners: a mixed-methods systematic review": S2 File

**Quantitative data collection form (adapted from Cochrane data extraction form)^[1]^**

**General information**

| Article Endnote ID |
| --- |
| Study ID *(surname of first author and year first full report of study was published e.g. Smith 2001)* |
| Article title |
| Year of publication |
| Year of study |
| Study location (country) |
| Study author contact details |

**Characteristics of included studies**

**Methods**

|  | **Descriptions as stated in report/paper** |
| --- | --- |
| Aim of study |  |
| Study design |  |
| Ethical approval obtained for study | ☐ ☐ ☐ ☐  Yes No Unclear Not applicable |

**Participants**

|  | Description  *(Include comparative information for each exposure or comparison group if available)* |
| --- | --- |
| Population description *(from which study participants are drawn)* |  |
| Site and Setting *(including location(s) and type of setting e.g. multicentre cohort study at the tertiary hospitals in Victoria State)* |  |
| Eligibility criteria for participants *(inclusion and exclusion criteria)* |  |
| Method of recruitment of participants, if applicable *(e.g. phone, mail, clinic patients)* |  |
| Informed consent obtained | ☐ ☐ ☐ ☐  Yes No Unclear Not applicable |
| Sample size (*e.g. N=30, F=20, M10)* |  |
| Response / participation rate, if applicable *(e.g. 80% responded)* |  |
| Age range or mean age, if applicable |  |
| Other relevant sociodemographic, if applicable |  |
| Subgroups, if applicable |  |

**Exposed groups**

**Exposed group 1 (Had PPH)**

|  | **Description as stated in report/paper** |
| --- | --- |
| Group name *(if more than one exposed group)* |  |
| Definition of exposure and data source |  |
| Definition of comparison group *(if any)* |  |
| Interventions received for PPH *(if applicable as exposed group criteria)* |  |
| Other Co-morbidities or pregnancy complications *(if applicable)* |  |

**Outcomes**

*(Copy and paste table for each outcome.)*

**Outcome 1**

|  | **Description as stated in report/paper** |
| --- | --- |
| Outcome name |  |
| Outcome definition and categories |  |
| Outcome measurement *(e.g. self-report, clinical diagnosis, records)* |  |
| Is outcome/tool validated? | ☐ ☐ ☐ ☐  Yes No Unclear Not applicable |
| Frequency and time of measurement *(in relation to birth, e.g. 1 month postpartum and 3 month postpartum)* |  |

**Other**

| Study funding sources *(including role of funders)* |
| --- |
| Possible conflicts of interest *(for study authors)* |
| Authors’ conclusions or recommendations as reported in the paper |
| Notes: |

**Risk of bias assessment (RoBANs tool)**

| **Domain** | **Risk of bias** | | | **Support for judgement**  *(include direct quotes where available with explanatory comments)* | **Location in text or source** *(pg & ¶/fig/table/other****)*** |
| --- | --- | --- | --- | --- | --- |
|  | **Low** | **High** | **Unclear** |  |  |
| 1. Participant selection | ☐ | ☐ | ☐ |  |  |
| 2. Confounding variables | ☐ | ☐ | ☐ |  |  |
| 3. Exposure measurement | ☐ | ☐ | ☐ |  |  |
| 4. Blinding and outcome measurement *(detection bias)* | ☐ | ☐ | ☐ | Outcome group: All/ |  |
| *(if separate judgement by outcome(s) required)* | ☐ | ☐ | ☐ | Outcome group: |  |
| 5. Incomplete data | ☐ | ☐ | ☐ |  |  |
| 6. Selective reporting | ☐ | ☐ | ☐ |  |  |
| Notes: | | | | | |

**Data and analysis**

*(Copy and paste the appropriate table for each outcome, including additional tables for each time point and subgroup as required.)*

**Dichotomous outcome**

|  | **Description as stated in report/paper** | | | |
| --- | --- | --- | --- | --- |
| Outcome |  | | | |
| Subgroup |  | | | |
| Time point(s) |  | | | |
| Results (Cohort study) | Exposed group (e.g. PPH) | | Unexposed group (e.g. No PPH) | |
|  | No. with event (n/ %) | Total in group | No. with event (n/%) | Total in group |
| Results (case control study) | Cases | | Control | |
|  | No. with PPH (n/%) | Total in group | No. without PPH (n/%) | Total in groups |
| Any other results reported *(e.g. odds ratio, risk difference, CI or P value)* |  | | | |
| Statistical methods used and appropriateness of these *(e.g. adjustment for correlation)* |  | | | |

**Continuous outcome**

|  | | **Description as stated in report/paper** | | | | |
| --- | --- | --- | --- | --- | --- | --- |
| Outcome | |  | | | | |
| Subgroup | |  | | | | |
| Time point(s) | |  | | | | |
| Results | Exposed | | | Non-exposed | | |
|  | Mean | SD *(or other variance, specify)* | No. participants | Mean | SD *(or other variance, specify)* | No. participants |
| Any other results reported *(e.g. mean difference, CI, P value)* | |  | | | | |
| Statistical methods used and appropriateness of these *(e.g. adjustment for correlation)* | |  | | | | |

**Other outcomes**

|  | **Description as stated in report/paper** | | | |
| --- | --- | --- | --- | --- |
| Comparison |  | | | |
| Outcome |  | | | |
| Subgroup |  | | | |
| Time point (s) |  | | | |
| No. participants | Exposed (e.g. PPH) (n/%) | | Unexposed (e.g. No PPH) (n/%) | |
| Results | Exposed result | SE (or other variance) | Control result | SE (or other variance) |
|  | Overall results | | SE (or other variance) | |
| Any other results reported |  | | | |
| Statistical methods used and adjusted confounders |  | | | |

Qualitative data collection form

**General Information**

| Article Endnote ID |
| --- |
| Study ID *(surname of first author and year first full report of study was published e.g. Smith 2001)* |
| Article title |
| Year of publication |
| Year of study |
| Study location *(country)* |
| Study author contact details |

**Characteristics of included studies**

**Methods**

|  | **Descriptions as stated in report/paper** |
| --- | --- |
| Aim of study |  |
| Study Design |  |
| Ethical approval needed/ obtained for study | ☐ ☐ ☐ ☐  Yes No Unclear Not applicable |

**Participants**

|  | **Descriptions as stated in report/paper** |
| --- | --- |
| Population description *(from which study participants are drawn)* |  |
| Site and Setting *(including location and social context)* |  |
| Eligibility criteria for participants *(inclusion and exclusion criteria)* |  |
| Sampling Method or Strategy for qualitative study *(e.g. snowball sampling, purposive sampling, convenience sampling, etc)* |  |
| Method of recruitment of participants, if applicable *(e.g. phone, mail, clinic patients)* |  |
| Informed consent obtained | ☐ ☐ ☐  Yes No Unclear/unreported |
| Sample size (*e.g. N=30, F=20, M10)* |  |
| Participants response/participation rate *(if applicable)* |  |
| Age range or mean age if applicable |  |
| Other relevant sociodemographic characteristics of participants *(if applicable)* |  |

**Data collection and analysis**

|  | **Description as stated in report/paper** |
| --- | --- |
| Data Collection | Summarise the data collection approach. Include: type of data collection/source (e.g. semi-structured interviews/FGDs), total number of interviews/Focused Group Discussions (FGDs)/ open-ended survey; how the data were collected (e.g. face to face/telephone/email) ; location (s) (e.g. home, clinics); number of participants in FGDs if applicable; Duration of interviews/FGDs (e.g. 60 to 90 mins); information for interviewers/facilitators (e.g. by an experienced midwives) ; timing of interviews (e.g. 3- 6 months postpartum); whether they are interviewed separately if the partners were included. |
| Questions or topics covered for interviews/ FGDs/Survey(s) | Describe topics covered by interviews/ FGD(s) and/or open-ended survey questions asked |
| Data Analysis *(Summarise approach to data analysis and synthesis, e.g. content analysis, thematic analysis, etc)*  Describe the theoretical approach if reported *(e.g. naturalistic inquiry, grounded theory)* |  |
| Results *(copy/paste the themes reported)* |  |

**Other**

| Study funding sources *(including role of funders)* |
| --- |
| Possible conflicts of interest *(for study authors)* |
| Authors’ Conclusion and Recommendation *(if applicable)* |
| Notes: |

1. Cochrane data collection form for intervention reviews: RCTs and non-RCTs. Version 3, April 2014. <https://dplp.cochrane.org/data-extraction-forms>
